# The FEES Dysphagia Index: a bias-resilient continuous score that captures expert clinical judgment in 2,943 neurological inpatients

**DOI:** 10.64898/2026.04.20.26351259

**Authors:** Cornelius J. Werner, Elizabet Sanchez-Garcia, Bettina Mall, Tareq Meyer, João Pinho, Joerg B. Schulz, Beate Schumann-Werner

## Abstract

Multi-consistency testing during flexible endoscopic evaluation of swallowing (FEES) is clinically necessary but introduces selection bias: worst scores inflate severity because the number of consistencies tested covaries with disease severity. In this retrospective observational study of hospitalized neurological patients, we derived and validated the FEES Dysphagia Index (FDI) in two temporally independent cohorts (Cohort 1: 2013-2018, N=1,257; Cohort 2: 2021-2025, N=1,686) from a single center. FDI-S averages Penetration-Aspiration Scale (PAS) scores across tested consistencies (0-100 scale); FDI-E uses Yale Pharyngeal Residue scores; FDI-C combines both. Selection bias was quantified using sequential branching-tree inverse probability weighting (IPW). Worst PAS overestimated severity by 24%; FDI deviated by <2%. FDI-C was significantly superior to Worst PAS for hospital-acquired pneumonia (HAP; AUC 0.70 vs. 0.60, p<0.001), mortality (0.71 vs. 0.62, p=0.040), and restricted oral intake (0.90 vs. 0.74, p<0.001), and statistically equivalent to clinician-rated severity. FDI-C mapped linearly onto ordinal Functional Oral Intake Scale values (FOIS; proportional odds RCS p=0.99). With functional status and diagnosis, FDI-C reconstructed the clinician’s oral intake recommendation with AUC up to 0.93. The FDI-C-mortality relationship was sigmoidal with a clinically relevant transition zone between ∼50 and ∼85. FDI-C is a bias-resilient, bedside-calculable score with interval-scale properties that captures expert clinical judgment, suitable as both a clinical decision support tool and a continuous research endpoint.

## Introduction

Flexible endoscopic evaluation of swallowing (FEES) is a cornerstone of dysphagia assessment in neurological patients. Since its introduction by Langmore and colleagues [1], FEES has evolved into a widely available, bedside-compatible procedure that allows direct visualization of pharyngeal swallowing physiology without radiation exposure [2]. Current guidelines endorse FEES as a first-line instrumental assessment in acute stroke, neurodegenerative disease, and critical care settings [3–5].

The Penetration-Aspiration Scale (PAS) [6] remains the most widely used outcome measure derived from FEES. However, both its psychometric properties and its application in research remain contentious. Steele and Grace-Martin [7] demonstrated that the PAS does not function as a truly ordinal scale, with non-equidistant intervals and rarely observed levels, and proposed a physiologically motivated reclassification. In a comprehensive systematic review, Borders and Brates [8] documented that 183 studies using the PAS employed at least ten different categorization schemes, with no consensus on whether the scale should be treated as ordinal, categorical, or interval. Critically, among studies that administered multiple bolus types or consistencies, 55% reported PAS results only at the participant or group level, typically as a worst score, discarding information about consistency-specific patterns.

In routine practice, clinicians adapt consistency selection to the patient’s presentation, omitting consistencies deemed unsafe or unnecessary - a process we term "clinical gating", in analogy to adaptive testing protocols in psychometrics where subsequent items are administered conditional on prior responses. Multi-consistency testing is clinically necessary, as bolus consistency is a primary determinant of penetration-aspiration risk: Borders and Steele [9], using Bayesian multilevel ordinal regression on two large videofluoroscopic datasets (n=678 and n=177, totaling over 11,500 bolus-level observations), demonstrated that thin liquids carry substantially higher PAS scores than thicker consistencies. However, multi-consistency testing introduces a methodological problem that has received little formal attention so far: number and type of consistencies tested will necessarily covary with disease severity, and therefore any summary statistic computed over a variable number of observations is biased when the number of observations depends on the outcome. Additionally, a survey of German FEES centers confirmed substantial variation in which consistencies are administered in the first place, further compounding this problem at a higher level in terms of comparing outcomes across different centers [10].

The consequences are well documented but unresolved. Borders and Steele [9] stated explicitly that "the common practice of summarizing PAS scores based on worst scores is a bias." An additional Monte Carlo simulation by Borders and colleagues [11] demonstrated that worst-score aggregation of PAS data reduces statistical power and inflates effect size estimates compared to multilevel approaches that use all available bolus trials. Yet no study has quantified the magnitude of this bias in the context of variable consistency testing of actual patients, and no scoring method has been proposed that addresses it.

The same problem affects existing composite scores. The Dynamic Imaging Grade of Swallowing Toxicity adapted for FEES (DIGEST-FEES) [12,13] uses the maximum PAS as its entry point for safety grading and the maximum pharyngeal residue for efficiency grading. The Fiberoptic Endoscopic Dysphagia Severity Scale (FEDSS) [14,15] assigns severity based on the worst finding across a sequential testing protocol. The ordinal FEES dysphagia score [16], a 4-level classification grading dysphagia from none through mild (premature spillage and/or residue without penetration/aspiration) to severe (penetration/aspiration of two or more consistencies), was proposed in the context of the integrated FEES report [4] and has been applied in studies of deep brain stimulation, myasthenia gravis, and Guillain-Barré syndrome [17–19]; it, too, relies on worst-finding logic across consistencies. Neither score adjusts for the variable number of consistencies administered, and both therefore inherit the structural vulnerability to clinical gating.

The primary objective of this study is to derive and temporally validate the FEES Dysphagia Index (FDI), a bias-resilient continuous score that corrects for clinical gating in multi-consistency FEES testing. Specifically, we derive FDI-S in a 2013-2018 cohort (N=1,257) and replicate its bias resilience in an independent 2021-2025 cohort (N=1,686) from the same center. Secondary objectives are (i) to quantify the selection bias introduced by clinical gating using sequential inverse probability weighting as a formal reference; (ii) to extend the framework to swallowing efficiency (FDI-E, based on Yale Pharyngeal Residue scores [20]) and a combined composite (FDI-C); (iii) to evaluate whether FDI-C captures the swallowing physiology component of clinical decision-making and its complementarity with functional status and diagnostic context, including its correspondence with clinician-assigned oral intake levels; and (iv) to assess FDI-C’s suitability as both a clinical decision support tool and a continuous endpoint for dysphagia research.

## Methods

### Study design

This retrospective two-cohort observational study analyzed consecutive neurological inpatients who underwent FEES at the Department of Neurology, RWTH Aachen University Hospital. Although the analysis was designed retrospectively, the core clinical variables (PAS per consistency, Yale Pharyngeal Residue scores, FOIS, and FEES severity summaries) were documented prospectively at the time of examination as part of a standardized clinical FEES protocol, which limits information bias and preserves consistent variable definitions across the observation period. Reporting follows the STROBE guidelines for observational studies (Online Resource 8).

Cohort 1 (derivation) comprised all FEES patients from January 2013 to December 2018 (N=1,257). Cohort 2 (replication and extension) comprised all FEES patients from January 2021 to December 2025 (N=1,686 of 18,851 total inpatient episodes). The two cohorts do not overlap. The 2019-2020 period was excluded by design to avoid atypical referral patterns, infection-control-driven restrictions on FEES testing, and modified clinical workflows during the COVID-19 pandemic.

FEES was performed as part of routine clinical care according to a standardized protocol [2,4].

The study was approved by the ethics committee of RWTH Aachen University (EK 088/17) and was performed in accordance with the ethical standards laid down in the 1964 Declaration of Helsinki and its later amendments. Individual informed consent was waived by the ethics committee due to the retrospective design and the use of routinely collected clinical data.

### FEES protocol

FEES examinations followed a sequential multi-consistency protocol. Up to four consistencies were tested: puree, thin liquid, thickened liquid (if deemed necessary after liquid), and solid food, in this order. Consistency selection followed clinical judgment: examiners could omit or add consistencies based on the patient’s clinical presentation and observed swallowing performance (clinical gating). For each tested consistency, the Penetration-Aspiration Scale (PAS; 1-8) [6] was documented. In Cohort 2, the Yale Pharyngeal Residue Severity Rating Scale (1-5, separately for valleculae and pyriform sinuses) [20] was additionally documented as part of enhanced routine documentation.

### FDI construction

#### FDI-S (Safety)

The arithmetic mean of PAS scores across all tested consistencies, linearly rescaled to 0-100 (100 = no impairment):

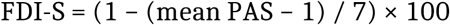

This formulation averages rather than maximizes, thereby normalizing for the variable number of consistencies tested. Unlike the Worst PAS, whose mathematically expected value increases with the number of observations, the arithmetic mean is unbiased with respect to the number of consistencies, provided the tested consistencies are representative of the untested ones conditional on observed covariates.

#### FDI-E (Efficiency; Cohort 2 only)

The arithmetic mean of Yale combined scores (mean of valleculae and pyriform sinus scores) across tested consistencies, rescaled to 0-100:

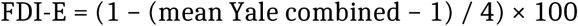

#### FDI composite (FDI-C; Cohort 2 only)

FDI-C = (FDI-S + FDI-E) / 2, giving equal weight to safety and efficiency. The equal-weight specification was chosen a priori to avoid outcome-specific optimization and to maximize clinical interpretability. A sensitivity analysis examining regression-derived and AUC-maximizing alternative weightings of FDI-S and FDI-E confirmed that equal weighting performs comparably across all outcomes (Online Resource 3). A parallel sensitivity analysis for the valleculae vs. pyriform sinus weighting within FDI-E confirmed that the simple Yale mean generalizes across consistencies better than regression-derived empirical weights, which reverse direction between consistencies (Online Resource 7).

### Comparator scores

#### Worst PAS

The maximum PAS across all tested consistencies [6,8].

#### Ordinal FEES severity score (Cohort 2)

A 4-level ordinal classification (0=none, 1=mild, 2=moderate, 3=severe) based on the combination of premature spillage, penetration-aspiration, and residue thresholds across tested consistencies [4,16]. This classification uses a maximum-based logic, assigning severity based on the worst findings.

#### Clinician-rated severity (Cohort 2)

A 7-level ordinal severity grading (0, 0.5, 1, 1.5, 2, 2.5, 3) with 0 = no dysphagia to 3 = severe dysphagia, extracted from the FEES summary text and converted to numerical values, capturing the clinician’s holistic severity judgment. Half-steps (e.g., 1.5 for "mild-to-moderate") accommodate composite gradings used in clinical practice.

### Clinical outcomes

#### Cohort 1

Hospital-acquired pneumonia (HAP), defined as ICD-10 J69 coded as any-position diagnosis during the inpatient stay and used as a pragmatic proxy for aspiration-related pneumonia given that J69 may also capture chemical pneumonitis (12.7%); in-hospital mortality (3.3%); and prolonged length of stay (>median 15 days, 48.8%; Online Resource 2).

#### Cohort 2

HAP (ICD-10 J69 as a secondary diagnosis, 14.9%), in-hospital mortality (4.3%), and restricted oral intake

### Inverse probability weighting

To quantify selection bias, we constructed inverse probability weights (IPW) using a sequential branching-tree model that mirrored the clinical testing decisions [22].

At each decision node (puree→liquid, liquid→solid, liquid→thickened liquid), a logistic propensity model estimated the probability of proceeding to the next consistency. Baseline covariates included age, sex, Hospital Frailty Risk Score (HFRS) [23], functional status (Self-Care Index at admission, SPI [24,25]), and neurological diagnosis. PAS scores from preceding consistencies were included at all branches.

In Cohort 2, the IPW specification was refined based on Cohort 1 experience: Yale residue scores were included as additional covariates only where likelihood ratio tests confirmed their predictive relevance (liquid→solid: p<0.001), and excluded from the liquid→thickened liquid branch a priori, as this "rescue" branch reflects a pure safety decision. Propensity scores were truncated at the 1st and 99th percentiles.

#### Cohort 1 IPW diagnostics

Stabilized weights had mean 1.00 (SD 0.87), median 0.61, range 0.20-5.45. No extreme-weight trimming was required.

#### Cohort 2 IPW diagnostics

Weights for the puree→liquid and liquid→solid branches were well-behaved (max 5.48). The thickened liquid branch showed unstable weights (max 39.3, 175 weights >10) due to a positivity violation at a relatively small 35% testing rate. A sensitivity analysis excluding this branch was performed. Branch-specific propensity model details, likelihood-ratio tests for Yale covariate inclusion, and stabilized weight distributions for both cohorts are reported in Online Resource 1.

### Bias quantification

We used a three-cohort design: (1) the complete-case subcohort (all four consistencies tested), (2) the standard-consistency subcohort (puree + liquid), and (3) the IPW-adjusted full cohort. The primary bias metric was the Naive-IPW Delta: |naive mean − IPW mean| / IPW mean × 100. A Naive-IPW Delta below 1% indicates that the naive FDI already approximates the causal estimand. Both the IPW framework and the FDI averaging approach assume sequential conditional independence (sequential missing-at-random / MAR): at each testing decision, the missingness of untested consistencies is independent of their potential outcomes, conditional on observed covariates including PAS and Yale scores from previously tested consistencies. Because the branching-tree model conditions each decision node on the strongest clinical predictor of the subsequent gating decision, namely the swallowing outcome observed at the preceding step, this assumption is substantially weaker than unconditional MAR. Additionally, the direction of any residual bias is predictable: if clinicians withhold testing when they expect worse outcomes beyond what the observed covariates capture, the IPW reference slightly underestimates true population severity, making the Naive-IPW Delta a conservative estimate of FDI’s bias resilience.

### Predictive validity

Discrimination was assessed using AUC with DeLong comparisons [26]. Significance was set at α=0.05 (two-sided). No correction for multiple comparisons was applied to DeLong tests, consistent with the exploratory nature of pairwise score comparisons; individual p-values should be interpreted accordingly.

### Scale properties (Cohort 2)

Existing FEES scores are ordinal at best: DIGEST-FEES yields 5 levels, FEDSS 6, and the ordinal FEES severity score 4. Ordinal scales limit the choice of statistical methods, reduce power in clinical trials [27,28], and discard within-category variation. Because FDI-C produces a quasi-continuous distribution by design, we assessed whether it meets the empirical criteria for interval-scale treatment. **Granularity:** Number of unique values per score. **Linearity of the logit:** Restricted cubic splines (RCS) with 4 knots [29]; here, a non-significant non-linearity test (p>0.05) supports interval-scale use. **Information loss:** AIC comparison of continuous versus quintile models. **Calibration:** Observed outcome rates across FDI-C deciles. **Construct validity against ordinal FOIS:** The relationship between FDI-C and the full ordinal FOIS scale (1-7) was assessed using Spearman correlation and proportional odds regression with RCS.

### Clinical decision model (Cohort 2)

In clinical practice, the FEES examiner determines swallowing severity and then contextualizes it within the patient’s functional status and clinical acuity to recommend an oral intake level (FOIS). A patient with moderate dysphagia but preserved independence and chronic adaptation will receive a more liberal diet than a patient with similar swallowing impairment who is acutely ill, frail, and lacks established protective mechanisms. We formalized this clinical reasoning in a statistical model:

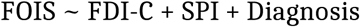

where FDI-C captures swallowing physiology, SPI captures functional status, and neurological diagnosis (stroke, neurodegenerative, other) serves as a proxy for acuity and chronicity. We fitted (A) a proportional odds model for ordinal FOIS (1-7), and (B) logistic models for binary FOIS≤3 and HAP, using nested model comparison (M1: FDI-C alone → M2: + SPI → M3: + Diagnosis). Incremental contributions were assessed by likelihood ratio tests and DeLong AUC comparisons.

For mortality (n=42 events in the complete-case subsample), the three-predictor model was not fitted because the events-per-variable ratio (EPV = 42/4 = 10.5) falls at the threshold below which logistic regression estimates become unreliable [30]. The FDI-C-mortality relationship was instead characterized descriptively using restricted cubic splines.

### Inter-rater reliability

Because FDI is computed deterministically from PAS and Yale ratings without additional rater-dependent judgments, its composite IRR was estimated using the Spearman-Brown prophecy formula [31]:

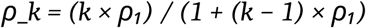

where ρ₁ is the single-consistency IRR and k is the number of consistencies tested. For FDI-S, we used ρ₁ = 0.85, representative of PAS IRR in FEES [8,32].

For FDI-E, we used ρ₁ = 0.751, based on the lowest published Yale Pharyngeal Residue IRR [20,33]. Because PAS IRR exceeds Yale IRR, the FDI-S component is expected to be at least as reliable as FDI-E, and the FDI-C composite reliability is bounded from below by the FDI-E estimate.

### Internal validation (Cohort 2)

FDI-S was validated by temporal replication across cohorts (see above). Because FDI-E and FDI-C are novel to Cohort 2, their predictive performance required independent confirmation. We assessed stability in a stratified 60/40 random split (derivation n=1,011, validation n=675), computing AUCs for all four outcomes in each split and comparing them using DeLong tests. Cross-split stability was defined as |ΔAUC| < 0.02 (stable), 0.02-0.05 (moderate), or > 0.05 (unstable). Additionally, the three-cohort bias analysis was repeated within each split to confirm that bias resilience was not dependent on the specific sample composition.

### Statistical analysis

All analyses were performed in R version 4.5.2 [34]. Key packages included pROC [35] for AUC analysis, rms [29] for restricted cubic splines, MASS for proportional odds regression, and cobalt for covariate balance diagnostics. Cohort 1 additionally used cumulative link mixed models (CLMMs) with the ordinal package for consistency-specific PAS analyses.

## Results

### Cohort characteristics

The combined sample comprised 2,943 neurological inpatients. Cohort 1 (N=1,257) had a mean age of 71.6 years (SD 13.7; median 74, IQR 64-82), was 55.0% male, and included 50.0% stroke, 22.3% neurodegenerative, and 27.7% other diagnoses. Hospital-acquired pneumonia (HAP) occurred in 160 patients (12.7%) and mortality in 42 (3.3%). Cohort 2 (N=1,686) included 57.1% stroke, 19.2% neurodegenerative, and 23.7% other diagnoses. HAP occurred in 251 (14.9%), mortality in 73 (4.3%), and restricted oral intake (FOIS≤3) in 392 (24.2%). Sixty-three patients in Cohort 2 (3.7%) had no valid FOIS documented and were treated as missing for FOIS-dependent analyses. Chart review identified three groups: failed or aborted examinations (n = 26), complete assessments with narrative documentation but missing structured fields (n = 24), and complete assessments with FDI scores where FOIS was simply not recorded (n = 13). None represented a distinct clinical state; the missing rate (3.7% overall, 0.8% among patients with calculable FDI) is unlikely to introduce systematic bias. Full characteristics are presented in Table 1.

**Table 1.** Patient characteristics.

| Characteristic | Cohort 1<br>(2013–2018, N = 1,257) | Cohort 2<br>(2021–2025, N = 1,686) |
| --- | --- | --- |
| Age, years, mean (SD) | 71.6 (13.7) | 70.6 (13.7) |
| Male sex, n (%) | 691 (55.0) | 950 (56.3) |
| **Neurological diagnosis** |  |  |
| Stroke | 629 (50.0) | 963 (57.1) |
| Neurodegenerative disease | 280 (22.3) | 324 (19.2) |
| Other neurological | 348 (27.7) | 399 (23.7) |
| **Functional status** |  |  |
| SPI at admission, mean (SD) | 26.0 (10.5)<sup>a</sup> | 27.7 (9.4) |
| HFRS, mean (SD) | 11.0 (6.2) | 11.4 (6.6) |
| **Outcomes** |  |  |
| HAP (J69) | 160 (12.7) | 251 (14.9) |
| In-hospital mortality | 42 (3.3) | 73 (4.3) |
| FOIS ≤ 3 | n/a | 392 (24.2)<sup>b</sup> |
| Prolonged LOS<sup>c</sup> | 613 (48.8) | 836 (49.6) |
| LOS, median (IQR), days | 15 (9–21) | 16 (8–23) |
| <sup>a</sup> n = 1,125 with SPI available. <sup>b</sup> Excludes 63 patients with missing/invalid FOIS. <sup>c</sup> > median (Cohort 1: > 15 d; Cohort 2: > 16 d).<br>LOS in Online Resource 2. |  |  |

### Consistency testing patterns

Both cohorts showed the expected clinical gating pattern. In Cohort 1, puree and thin liquid were tested in >91% of patients, whereas thickened liquid and solid were tested in 43.8% and 63.4%, respectively; only 331 patients (26.3%) received all four consistencies. In Cohort 2, testing rates were similar: puree 91.3%, liquid 89.0%, solid 68.0%, thickened liquid 35.3%; complete cases n=412 (24.4%). The consistency of these patterns across two cohorts separated by a temporal gap confirms that clinical gating is a stable institutional practice, not a sampling artifact.

### Consistency-specific safety profiles (Cohort 1)

The aspiration risk hierarchy was consistent across cohorts. In Cohort 1, thin liquid carried the highest risk (mean PAS 4.33; 36.0% aspiration), followed by thickened liquid (3.71; 26.2%), puree (2.26; 11.3%), and solid (1.36; 1.9%). In CLMMs, thin liquid (β=2.087, p<0.001) and thickened liquid (β=1.186, p<0.001) were associated with worse PAS relative to puree, while solid was associated with better scores (β=−1.339, p<0.001); these effects were robust to adjustment for age, sex, HFRS, and SPI. Cohort 2 confirmed this hierarchy: thin liquid (mean PAS 4.77; 48.0% aspiration) > thickened liquid (3.52; 20.8%) > puree (1.93; 7.7%) > solid (1.16; 1.2%). The hierarchy was unchanged after IPW adjustment in Cohort 1, confirming that it reflects genuine consistency-specific risk rather than a selection artifact.

Inter-consistency PAS correlations were moderate in both cohorts (Cohort 1: Spearman ρ=0.14-0.33; Cohort 2: ρ=0.13-0.33), confirming that swallowing safety with one consistency does not reliably predict safety with others and providing empirical support for multi-consistency testing. Yale residue correlations in Cohort 2 were higher (ρ=0.38-0.60), consistent with pharyngeal residue being a more stable trait across consistencies. Full pairwise correlation matrices for PAS and Yale across both cohorts are provided in Online Resource 5.

### Bias resilience

#### Cohort 1 (derivation)

Worst PAS was inflated by 24.1% in the complete-case subcohort (i.e., patients tested on all four consistencies; mean 6.28 vs. IPW reference 5.06) and underestimated by 10.4% in the standard-consistency subcohort (restricted to puree and liquid only; mean 4.53). FDI-S deviated by <2% from the IPW reference in both subcohorts (complete case: 67.15 vs. 68.43, bias +1.9%; standard consistency: 67.77 vs. 68.43, bias +1.0%). IPW correction changed FDI-S AUCs by <0.01.

#### Cohort 2 (replication and extension)

The Naive-IPW Delta was 0.5% for FDI-S, 0.6% for FDI-E, and 0.6% for FDI-C. Worst PAS showed a complete-case bias of 9.8% (complete-case mean 7.02 vs. IPW 6.39). This illustrates a well-known property of order statistics: the expected value of the maximum of a set of random variables increases with the number of observations, even when the underlying distribution is unchanged [36]. The complete-case subcohort consists of patients who were mild enough to proceed through all four testing stages (confirmed by their lower FDI-S), yet their Worst PAS is paradoxically inflated because having four rather than two or three tested consistencies provides more opportunities for an extreme PAS value on any single trial.

We performed a sensitivity analysis to test whether FDI’s bias resilience depends on the specific IPW model. The thickened liquid branch had unstable weights (positivity violation at 35% testing rate), so we re-ran the IPW excluding this branch entirely. This changed the IPW reference values substantially (e.g., FDI-S shifted by 8.4 points between the two IPW specifications), indicating that the exact IPW-corrected population mean is sensitive to modeling choices. However, the Naive-IPW Delta - the discrepancy between the simple, unweighted FDI mean and the IPW-corrected mean - remained below 1% under both specifications. This means that regardless of which IPW model is used as the reference, the naive FDI consistently lands close to it. The practical implication is that FDI does not require IPW correction: the simple bedside-calculated mean is already a good approximation of the bias-corrected value. Detailed three-cohort bias results under both IPW specifications (full model and excluding the thickened liquid branch) are reported in Online Resource 6.

Results are presented in Table 3 and Figure 1.

**Fig. 1.**
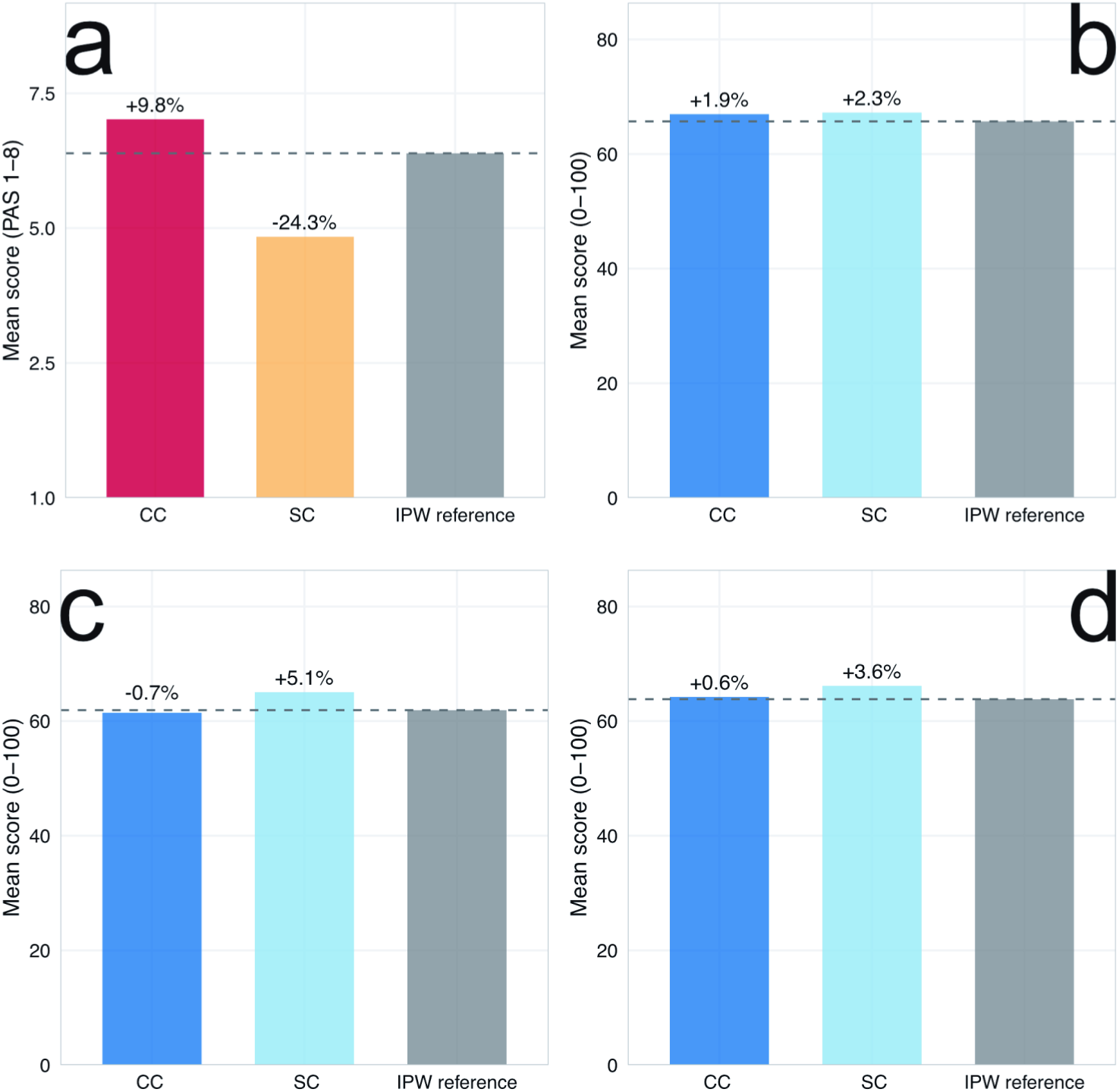
Three-cohort bias comparison for Cohort 2. Mean scores for the complete-case subcohort (all four consistencies tested, n = 412), the standard-consistency subcohort (puree + liquid, n = 1,480), and the IPW-adjusted reference (n = 1,559). **a** Worst PAS (1-8 scale). **b-d** FDI variants (0-100 scale), with b = FDI-S, c = FDI-E and d = FDI-C. Percentage labels indicate complete-case (CC) bias and standard-consistency (SC) bias relative to the IPW reference

**Table 2.** Consistency testing patterns and FDI score distributions.

| Panel A: Consistencies tested |  |  |  |  |  |
| --- | --- | --- | --- | --- | --- |
| Consistency |  | Cohort 1 (N = 1,257) |  | Cohort 2 (N = 1,686) |  |
| Puree |  | 1,145 (91.1) |  | 1,539 (91.3) |  |
| Thin liquid |  | 1,156 (92.0) |  | 1,500 (89.0) |  |
| Solid |  | 797 (63.4) |  | 1,147 (68.0) |  |
| Thickened liquid |  | 550 (43.8) |  | 595 (35.3) |  |
| Panel B: Testing pathways |  |  |  |  |  |
| No. of consistencies |  | Cohort 1 |  | Cohort 2 |  |
| 1 |  | 113 (9.0) |  | 51 (3.0) |  |
| 2 |  | 228 (18.1) |  | 175 (10.4) |  |
| 3 |  | 585 (46.5) |  | 893 (53.0) |  |
| 4 (all) |  | 331 (26.3) |  | 412 (24.4) |  |
| Panel C: Score distributions (Cohort 2) |  |  |  |  |  |
| Score | n | Mean (SD) | Median | Range | Unique values |
| FDI-S | 1,559 | 72.6 (25.7) | 75.0 | 0–100 | 40 |
| FDI-E | 1,557 | 64.5 (21.3) | 66.7 | 0–100 | 45 |
| FDI-C | 1,557 | 68.6 (20.6) | 71.1 | 0–100 | 387 |
| Worst PAS | 1,559 | 4.9 (2.8) | 5.0 | 1–8 | 8 |
| Ordinal FEES severity | 1,686 | 2.1 (0.9) | — | 0–3 | 4 |
| Clinician-rated severity | 1,535 | 1.7 (0.9) | — | 0–3 | 7 |
| Panel D: Score distributions (Cohort 1, FDI-S only) |  |  |  |  |  |
| Score | n | Mean (SD) | Median | Range | Unique values |
| FDI-S | 1,257 | 70.2 (30.6) | 75.0 | 0–100 | 41 |
| Worst PAS | 1,257 | 4.6 (2.8) | 5.0 | 1–8 | 8 |

**Table 3.** Bias resilience: three-cohort analysis.

| Panel A: Cohort 1 (derivation) |  |  |  |  |  |
| --- | --- | --- | --- | --- | --- |
| Score | CC mean<br>(n = 331) | SCD mean<br>(n = 1,062) | IPW ref<br>(N = 1,257) | CC bias (%) | SCD bias (%) |
| Worst PAS | 6.28 | 4.53 | 5.06 | +24.1 | -10.4 |
| FDI-S | 67.15 | 67.77 | 68.43 | -1.9 | -1.0 |
| IPW diagnostics: mean 1.00 (SD 0.87), median 0.61, range 0.20–5.45. |  |  |  |  |  |
| Panel B: Cohort 2 (replication) |  |  |  |  |  |
| Score | CC mean<br>(n = 412) | SCD mean<br>(n = 1,480) | IPW ref<br>(n = 1,559) | CC bias (%) | Naive-IPW Δ<br>(%) |
| Worst PAS | 7.02 | 4.84 | 6.39 | +9.8 | n/a |
| FDI-S | 66.97 | 67.24 | 65.70 | +1.9 | 0.5 |
| FDI-E | 61.45 | 65.04 | 61.90 | -0.7 | 0.6 |
| FDI-C | 64.21 | 66.14 | 63.81 | +0.6 | 0.6 |
| CC = complete case; SCD = standard consistency design (puree + liquid); Naive-IPW Δ = \|naive - IPW\| / IPW × 100. |  |  |  |  |  |
| Panel C: Sensitivity (IPW without thickened liquid, Cohort 2) |  |  |  |  |  |
| Score | IPW ref (full) | IPW ref (no<br>thick) | Δ references | Naive-IPW Δ |  |
| FDI-S | 65.70 | 74.13 | +8.43 | 0.5 |  |
| FDI-C | 63.81 | 69.73 | +5.92 | 0.6 |  |

### Predictive validity

#### Cohort 1

FDI-S consistently outperformed Worst PAS: HAP (AUC 0.712 vs. 0.644, p<0.001), mortality (0.639 vs. 0.600, p=0.020), prolonged LOS (0.584 vs. 0.551, p<0.001).

#### Cohort 2

FDI-C was significantly superior to Worst PAS for all outcomes: HAP (0.698 vs. 0.604, ΔAUC=+0.094, p<0.001), mortality (0.706 vs. 0.622, +0.083, p=0.040), and FOIS≤3 (0.900 vs. 0.735, +0.164, p<0.001). FDI-C was also significantly superior to the ordinal FEES severity score for HAP (p=0.001) and FOIS≤3 (p<0.001).

FDI-C was statistically equivalent to clinician-rated severity for all outcomes (all p≥0.148). For HAP, FDI-C achieved AUC 0.698 versus 0.707 for clinician-rated severity (p=0.731). For FOIS≤3, clinician-rated severity reached 0.916, numerically exceeding FDI-C (0.900), but the difference was not significant (p=0.201). FDI-C correlated more strongly with clinician-rated severity (ρ=−0.829) than the ordinal FEES severity score did (ρ=0.611 with clinician-rated severity), suggesting that a simple arithmetic mean captures the holistic assessment that experienced clinicians perform intuitively.

IPW correction changed AUC values by ≤0.008 for all FDI variants, confirming that bias resilience at the score level translates to the inferential level.

Results are presented in Table 4. Prolonged LOS results are reported in Online Resource 2.

**Table 4.** Predictive validity: AUC comparison.

| Panel A: Cohort 1 (FDI-S only) |  |  |  |
| --- | --- | --- | --- |
| Score | Asp. pneumonia | Mortality |  |
| FDI-S | 0.712 (0.671–0.754) | 0.639 (0.553–0.724) |  |
| Worst PAS | 0.644 (0.604–0.683) | 0.600 (0.521–0.679) |  |
| DeLong (FDI-S vs Worst PAS) | +0.069, $p < 0.001$ | +0.039, $p = 0.020$ | |
| $N = 1,257$. Prolonged LOS: FDI-S $\rho = -0.200$ vs Worst PAS $\rho = 0.131$. | | | |
| Panel B: Cohort 2 (extended comparators) |  |  |  |
| Score | Asp. pneumonia | Mortality | FOIS $\leq 3$ |
| FDI-C | 0.698 (0.661–0.736) | 0.706 (0.649–0.763) | 0.900 (0.881–0.919) |
| FDI-S | 0.690 (0.652–0.728) | 0.692 (0.634–0.750) | 0.899 (0.880–0.919) |
| FDI-E | 0.656 (0.618–0.695) | 0.672 (0.612–0.731) | 0.777 (0.749–0.804) |
| Worst PAS | 0.604 (0.569–0.640) | 0.622 (0.567–0.678) | 0.735 (0.710–0.760) |
| Ordinal FEES sev. | 0.611 (0.574–0.648) | 0.619 (0.558–0.680) | 0.721 (0.694–0.748) |
| Clinician-rated sev. | 0.707 (0.675–0.740) | 0.760 (0.713–0.807) | 0.916 (0.900–0.931) |
| Panel C: DeLong comparisons, FDI-C vs comparators (Cohort 2) |  |  |  |
| Comparison | Asp. pneumonia | Mortality | FOIS $\leq 3$ |
| vs Worst PAS | +0.094, $p < 0.001$ | +0.083, $p = 0.040$ | +0.164, $p < 0.001$ |
| vs Ordinal FEES sev. | +0.087, $p = 0.001$ | +0.087, $p = 0.042$ | +0.178, $p < 0.001$ |
| vs Clin. severity | –0.009, $p = 0.731$ | –0.055, $p = 0.148$ | –0.016, $p = 0.201$ |
| Values are AUC (95% CI). Prolonged LOS in Online Resource 2. |  |  |  |

### Scale properties

FDI-C produced 387 unique values (ratio 0.249) - 48 times more than Worst PAS (8 values, ratio 0.005) and nearly 100 times more than ordinal severity classifications. The number of unique FDI-C values increased with k (k=1: 31, k=2: 111, k=3: 196, k=4: 176), whereas Worst PAS naturally remained at 8 regardless of k.

For HAP, the logit relationship was linear (RCS non-linearity p=0.601), and continuous treatment was preferred over quintiles by AIC (ΔAIC=+7.6). For FOIS≤3, the relationship was borderline linear (p=0.062). Calibration across deciles showed monotonic gradients for HAP (rate 37.2% → 3.8%) and FOIS≤3 (90.2% → 1.6%).

FDI-C correlated strongly with ordinal FOIS (Spearman ρ=0.731, n=1,545). Mean FDI-C showed a monotonic gradient across all seven FOIS levels: 36.5 (FOIS 1, nothing by mouth), 42.4 (FOIS 2), 53.2 (FOIS 3), 54.3 (FOIS 4), 69.1 (FOIS 5, oral diet with special preparation), 74.4 (FOIS 6), and 87.2 (FOIS 7, total oral diet). In a proportional odds model, the FDI-C-FOIS relationship was perfectly linear (RCS non-linearity p=0.986) - the strongest linearity result in the entire analysis.

Results are presented in Table 6 and Figure 2.

**Fig. 2.**
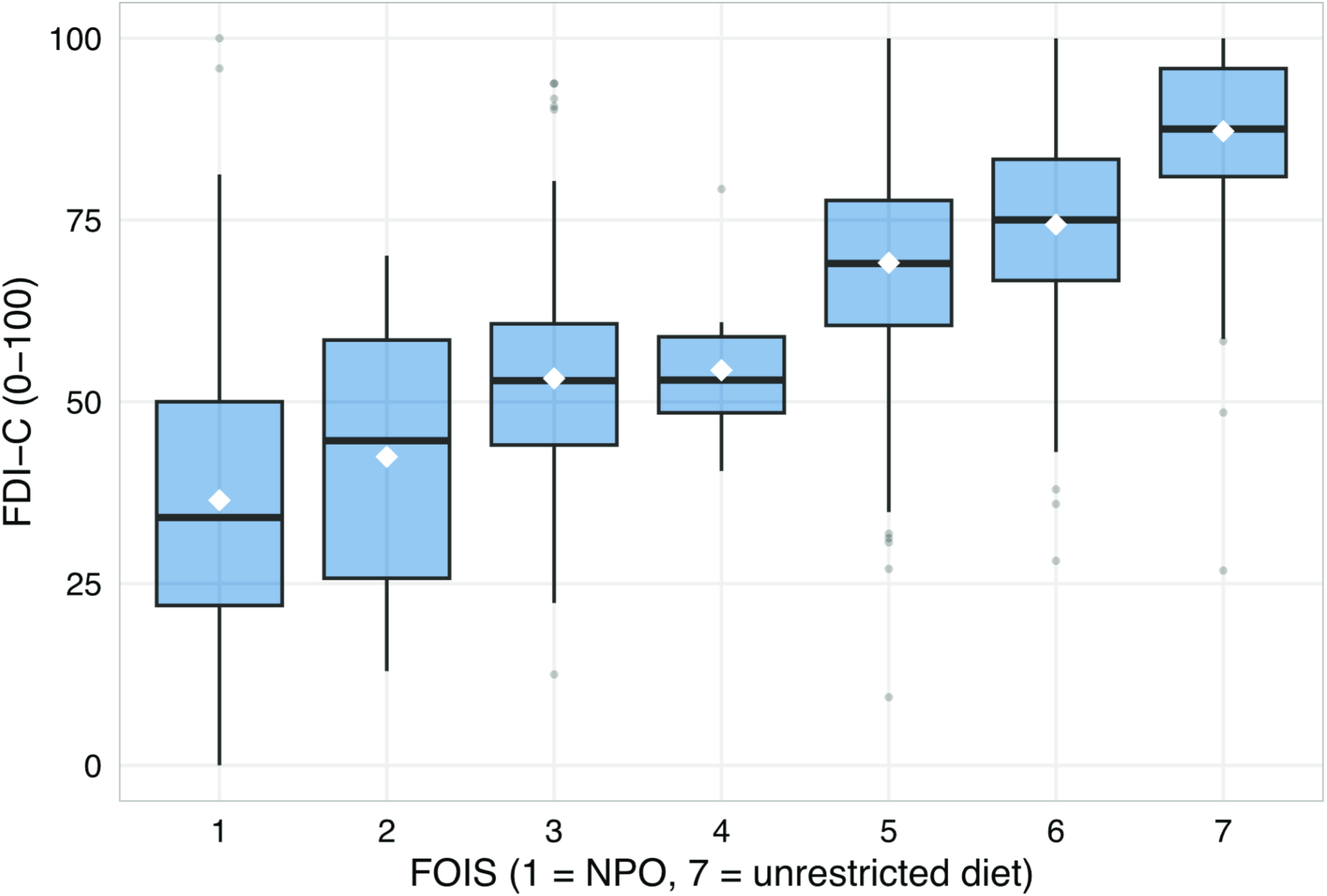
FDI-C and oral intake. Distribution of FDI-C by ordinal FOIS level (1-7) in Cohort 2. Boxes represent interquartile range, horizontal lines represent medians, whiskers extend to 1.5 times the interquartile range. Note: In this neurological inpatient cohort, FOIS = 4 (total oral intake of a single consistency) is rarely recommended; this pattern may differ in head-and-neck oncology cohorts, for which the FOIS was originally developed and single-consistency diets are more commonly prescribed

**Table 5.** Clinical decision model: FDI-C + SPI + Diagnosis (Cohort 2)

| Panel A: Model progression, AUC for binary outcomes |  |  |  |  |
| --- | --- | --- | --- | --- |
| Model | FOIS ≤ 3 (n = 1,376) |  | Asp. pneumonia (n = 1,386) |  |
| M1: FDI-C | 0.899 (0.878–0.920) |  | 0.688 (0.646–0.730) |  |
| M2: + SPI | 0.920 (0.902–0.938) |  | 0.799 (0.769–0.830) |  |
| M3: + Diagnosis | 0.930 (0.913–0.947) |  | 0.810 (0.782–0.839) |  |
| LR: SPI adds to M1 | χ<sup>2</sup> = 83.1, p < 0.001 |  | χ<sup>2</sup> = 119.1, p < 0.001 |  |
| LR: Dx adds to M2 | χ<sup>2</sup> = 39.5, p < 0.001 |  | χ<sup>2</sup> = 16.6, p < 0.001 |  |
| DeLong: M2 vs M1 | +0.021, p < 0.001 |  | +0.111, p < 0.001 |  |
| DeLong: M3 vs M2 | +0.010, p < 0.001 |  | +0.011, p = 0.009 |  |
| Panel B: Ordinal FOIS model (proportional odds, n = 1,376) |  |  |  |  |
| Model | AIC |  | McFadden R<sup>2</sup> |  |
| M1: FDI-C | 3,138.8 |  | 0.264 |  |
| M2: + SPI | 2,862.4 |  | 0.330 |  |
| M3: + Diagnosis | 2,727.2 |  | 0.362 |  |
| Prediction accuracy (M3): exact FOIS match 63.1%, within ±1 level 77.0%. |  |  |  |  |
| Panel C: Full model (M3) coefficients, FOIS ≤ 3 |  |  |  |  |
| Predictor | β | SE | OR | p |
| FDI-C | −0.1083 | 0.0071 | 0.897 | < 0.001 |
| SPI | −0.0777 | 0.0112 | 0.925 | < 0.001 |
| Dx: Neurodeg. vs Stroke | −2.1594 | 0.4161 | 0.115 | < 0.001 |
| Dx: Other vs Stroke | −0.6721 | 0.2334 | 0.511 | 0.004 |
| Panel D: FOIS ≤ 3 rate by FDI-C quartile and diagnosis |  |  |  |  |
| FDI-C quartile | Stroke | Neurodegenerative | Other |  |
| Q1 (worst) | 78.6% (n = 224) | 28.9% (n = 38) | 56.5% (n = 85) |  |
| Q2 | 18.7% (n = 203) | 0.0% (n = 60) | 10.0% (n = 80) |  |
| Q3 | 9.4% (n = 191) | 1.2% (n = 86) | 6.0% (n = 67) |  |
| Q4 (best) | 2.8% (n = 144) | 0.9% (n = 115) | 1.2% (n = 83) |  |
| Panel E: Full model (M3) coefficients, hospital-acquired pneumonia |  |  |  |  |
| Predictor | β | SE | OR | p |
| FDI-C | −0.0189 | 0.0040 | 0.981 | < 0.001 |
| SPI | −0.0994 | 0.0114 | 0.905 | < 0.001 |
| Dx: Neurodeg. vs Stroke | −1.4182 | 0.4097 | 0.242 | < 0.001 |
| Dx: Other vs Stroke | −0.2028 | 0.2135 | 0.816 | 0.342 |

**Table 6.**
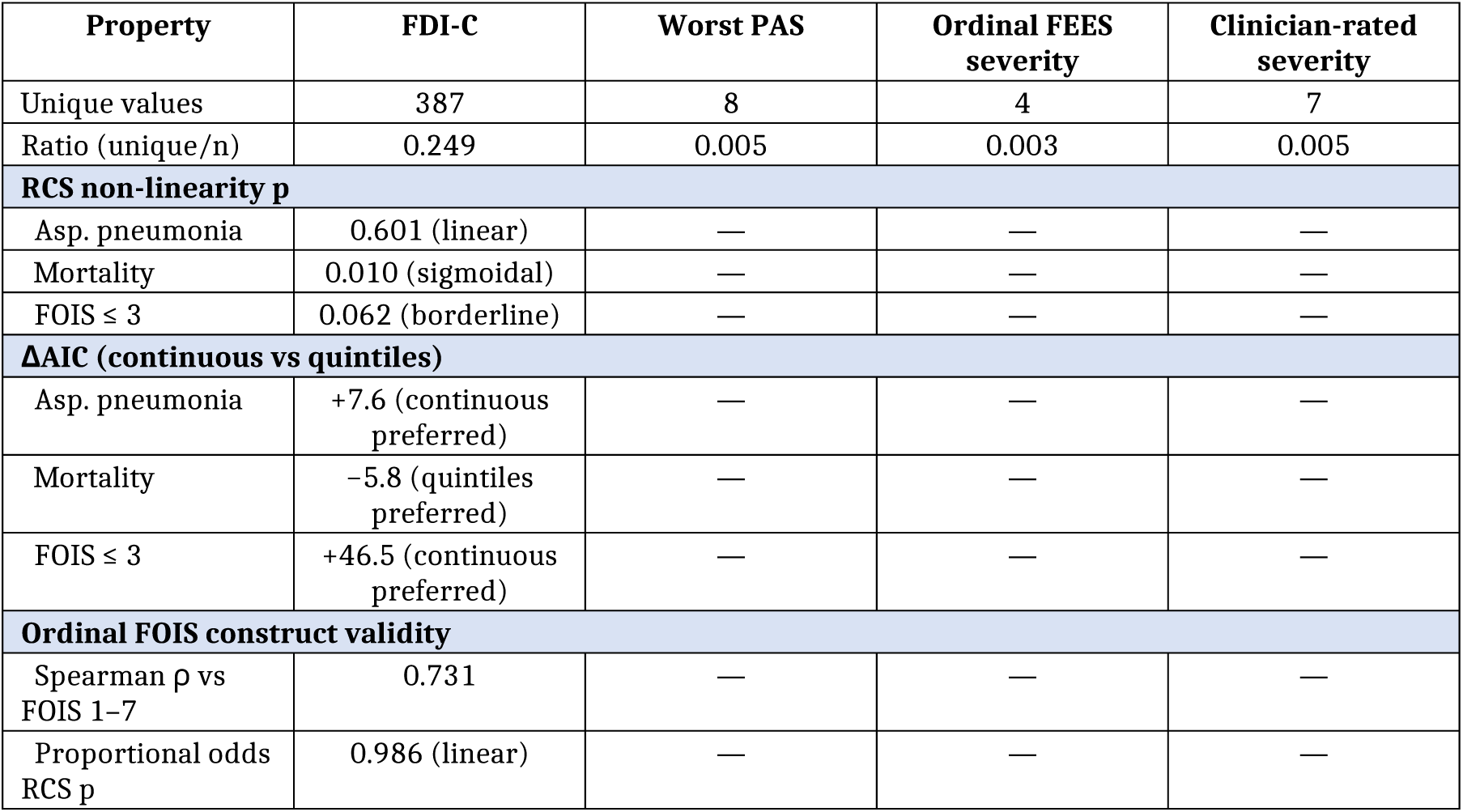
Scale properties of FDI-C (Cohort 2)

### FDI-C as the swallowing component of clinical decision-making

The clinical decision model confirmed that FDI-C, functional status, and diagnosis each contribute independently to predicting the clinician’s FOIS recommendation. In a proportional odds model for ordinal FOIS (n=1,376), all three predictors were highly significant (all p<0.001). Model fit improved progressively: AIC decreased from 3,138.8 (FDI-C alone, McFadden R²=0.264) to 2,862.4 (+ SPI, R²=0.330) to 2,727.2 (+ diagnosis, R²=0.362). The full model correctly predicted the exact FOIS level in 63.1% of cases and was within ±1 FOIS level in 77.0%.

Relative to stroke, neurodegenerative patients received more liberal diets at equivalent swallowing severity and functional status (β=+1.86, p<0.001), while "other" diagnoses showed a smaller effect in the same direction (β=+0.49, p<0.001). In the cross-tabulation of FDI-C quartiles by diagnosis, among patients with the worst swallowing (Q1), 78.6% of stroke patients received FOIS≤3 versus only 28.9% of neurodegenerative patients. In the best quartile (Q4), the groups converged (1-3% FOIS≤3).

For binary outcomes, the three-predictor model achieved AUC 0.930 for FOIS≤3 (vs. 0.899 for FDI-C alone, +0.031; each step significant at p<0.001) and AUC 0.810 for HAP (vs. 0.688 for FDI-C alone, +0.122). For HAP, the diagnosis effect was driven by neurodegenerative patients (OR=0.242, p<0.001); "other" diagnoses did not differ from stroke (OR=0.816, p=0.342).

### The mortality transition zone

The relationship between FDI-C and in-hospital mortality did not follow the linear pattern observed for HAP (RCS non-linearity p=0.010). The restricted cubic spline revealed an inverse sigmoidal curve with three approximate regions (Fig. 3), which should be interpreted as hypothesis-generating given the limited number of events (65 deaths): an approximate safe plateau above FDI-C ∼85 (mortality 0.7-1.1% across deciles D8-D10), an approximate transition zone between FDI-C ∼50 and ∼85 where mortality increased steeply from approximately 1% to 7-11%, and an approximate risk plateau below FDI-C ∼50 where mortality rates no longer increased further (D1: 7.1%, D2: 10.8%). Confidence intervals were wide, particularly in the tails (Fig. 3). 1

**Fig. 3.**
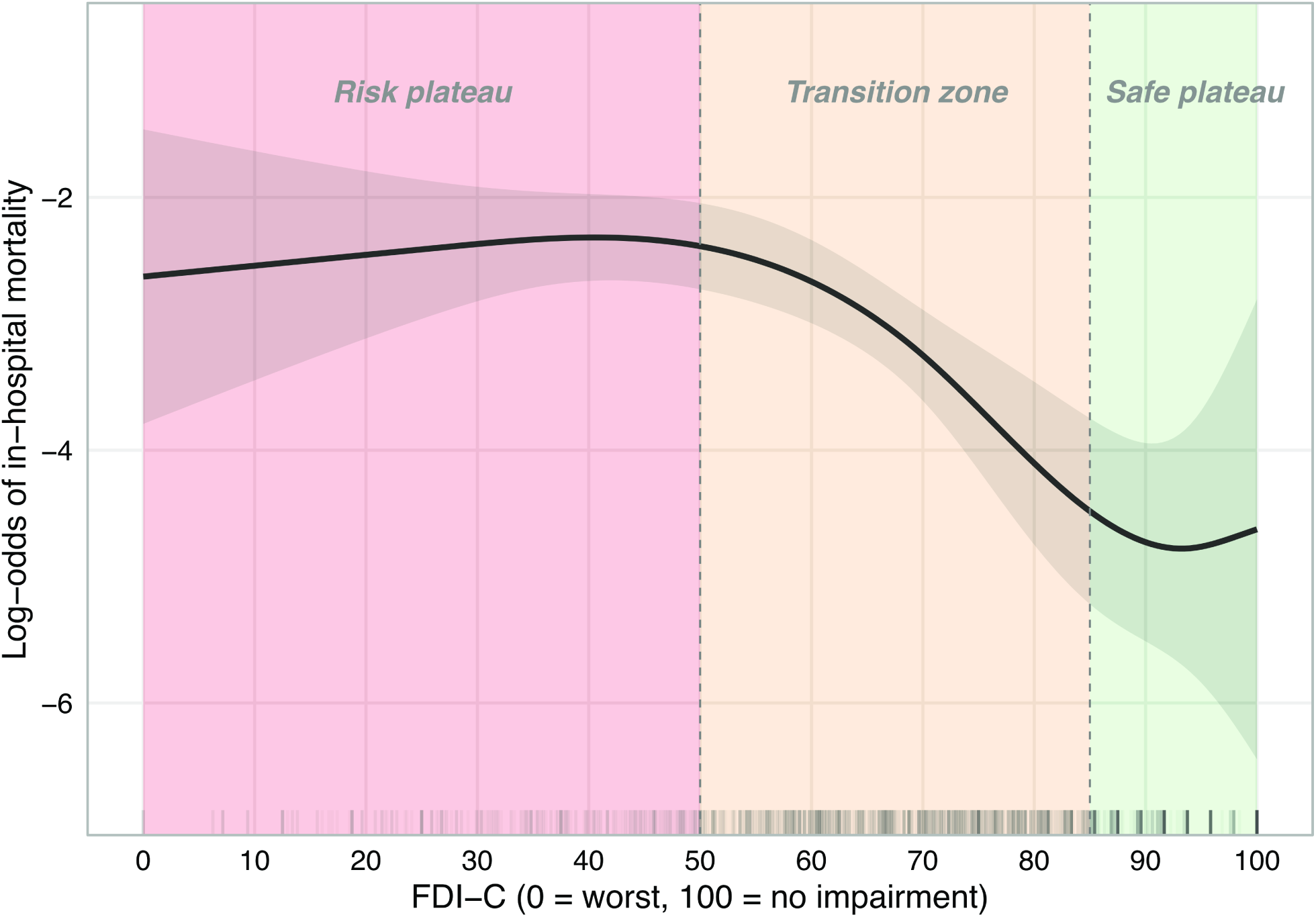
Restricted cubic spline (4 knots) showing the log-odds of in-hospital mortality as a function of FDI-C (Cohort 2, n = 1,557, 65 deaths; non-linearity p = 0.010). Shaded area represents the 95% confidence band. Shaded background regions indicate approximate zone assignments: risk plateau (FDI-C < 50), transition zone (50--85), safe plateau (> 85). Tick marks at the bottom represent individual patient FDI-C values

Results are presented in Table 7 and Figure 3.

**Table 7.** Mortality transition zone: calibration across FDI-C deciles (Cohort 2)

| Decile | n | Deaths | Mortality (%) | Mean FDI-C | Zone |
| --- | --- | --- | --- | --- | --- |
| D1 | 156 | 11 | 7.1 | 26.2 | Risk plateau |
| D2 | 157 | 17 | 10.8 | 47.7 | Risk plateau |
| D3 | 154 | 8 | 5.2 | 57.0 | Transition |
| D4 | 164 | 13 | 7.9 | 62.7 | Transition |
| D5 | 152 | 6 | 3.9 | 68.6 | Transition |
| D6 | 152 | 1 | 0.7 | 73.7 | Transition |
| D7 | 172 | 5 | 2.9 | 78.9 | Transition |
| D8 | 143 | 1 | 0.7 | 84.4 | Safe plateau |
| D9 | 177 | 2 | 1.1 | 90.7 | Safe plateau |
| D10 | 130 | 1 | 0.8 | 98.8 | Safe plateau |
| *Total: n = 1,557, 65 deaths (4.2%). Zone assignments approximate, based on RCS spline shape.* |  |  |  |  |  |

### Inter-rater reliability

FDI introduces no rater judgment beyond PAS and Yale, unlike DIGEST-FEES, which requires frequency classification, aspiration amount estimation, and residue percentage grading [12]. We estimated composite IRR using the Spearman-Brown formula rather than direct measurement, relying on published single-consistency IRR values from other centers [8,32,33]. By the Spearman-Brown formula, expected κ for FDI-S (based on PAS ρ₁=0.85) ranges from 0.92 (k=2) through 0.94 (k=3) to 0.96 (k=4), exceeding published DIGEST-FEESsafety IRR (κ_w=0.86 [12], 0.83 [13]) from k≥2 consistencies onward.

For FDI-E (based on Yale ρ₁=0.751), the corresponding estimates are 0.86 (k=2), 0.90 (k=3), and 0.92 (k=4), confirming that the efficiency component is also a reliable measure, again exceeding published DIGEST-FEES_efficiency_ IRR (κ_w=0.74 [12], 0.80 [13]).

The FDI-C composite reliability is thus bounded from below by the FDI-E estimates. We thus estimate the IRR of FDI-C to be κ≥0.86 at a minimum, with κ≥0.90 for the 84% of patients tested on three or more consistencies. This compares favorably with published DIGEST-FEES IRR (κ_w=0.83 [12], 0.82 [13]).

### Internal validation

In the 60/40 random split, FDI-C was stable: HAP AUC derivation 0.709 vs. validation 0.687 (Δ=−0.022); FOIS≤3 0.902 vs. 0.896 (Δ=−0.006); mortality 0.703 vs. 0.709 (Δ=+0.006). FDI-C remained significantly superior to Worst PAS in the validation set for HAP (p=0.007) and FOIS≤3 (p<0.001), and equivalent to clinician-rated severity (all p≥0.243). Full split AUCs for FDI-S, FDI-E, and FDI-C across all outcomes are provided in Online Resource 4.

## Discussion

### Principal findings

This study establishes the FEES Dysphagia Index through four stages: (1) identification and quantification of selection bias from clinical gating in multi-consistency FEES testing, (2) derivation of FDI-S as a bias-resilient alternative (Cohort 1, N=1,257), (3) temporal replication of bias resilience and extension to efficiency (FDI-E) and a composite (FDI-C) in Cohort 2 (N=1,686), and (4) demonstration that FDI-C captures the swallowing physiology component of clinical decision-making and has properties suitable for both clinical use and dysphagia research. Four findings merit detailed discussion.

### Selection bias: quantifying a known but unaddressed problem

The selection bias introduced by clinical gating has been recognized qualitatively [7–9,11], but no study has quantified its magnitude in the context of variable consistency testing.

Our data extend these observations by providing the first formal quantification in the context of variable consistency testing. In Cohort 1, Worst PAS was inflated by 24% in complete-case analyses. In Cohort 2, the bias was 10%. The difference reflects cohort composition (Cohort 1 had higher thickened liquid testing rates), but the pattern is consistent: maximum-based scores are systematically biased by clinical gating. That FDI-S deviated by <2% from the IPW reference in Cohort 1 and <1% in Cohort 2 demonstrates that simple averaging inherently corrects for this bias.

The mechanism is straightforward. By dividing by the number of consistencies tested, the arithmetic mean normalizes for the variable denominator that inflates maximum statistics. The IPW analysis confirms that this correction approximates a formal causal estimand, i.e., the score that would be observed under complete testing. That a bedside-calculable arithmetic mean closely approximates the output of a formal causal inference framework without requiring any covariate information is an unexpected finding, but underscores the practical advantage of the averaging approach.

The bias resilience of FDI rests on a sequential conditional independence assumption: for each untested consistency, its potential score is independent of the testing decision, conditional on scores from previously tested consistencies and patient covariates. This is weaker than the unconditional missing-at-random (MAR) assumption because the branching-tree IPW model conditions each decision node on the strongest predictor of the gating decision: the observed PAS at the preceding step. Under a strict missing-not-at-random (MNAR) scenario, where clinicians possess additional severity information not already captured by PAS, Yale scores, demographics, frailty, and functional status, both FDI and the IPW reference could underestimate true population severity. However, three observations limit the plausible magnitude of such residual bias: (1) the propensity models showed good discrimination at each branch, (2) the Naive-IPW Delta was consistent across two independent cohorts with different testing rates and different IPW specifications, and (3) the direction of any MNAR bias is predictable (underestimation of severity), meaning that the reported Naive-IPW Delta is conservative. We therefore consider residual MNAR bias unlikely to meaningfully affect the conclusions, while acknowledging that it cannot be formally excluded.

This central finding of bias resilience has implications beyond FDI. As noted in the Introduction, all major FEES severity scores rely on maximum-based or worst-finding logic and are therefore susceptible to the same bias, while the FDI framework for all practical purposes is not. An empirical comparison between FDI and these systems would be informative but is beyond the scope of this study.

### FDI-C captures the swallowing component of clinical decision-making

The most clinically important finding is that FDI-C, a bedside-calculable arithmetic mean, captures a substantial portion of the swallowing physiology input to the clinical diet recommendation. Three lines of evidence support this interpretation.

First, FDI-C maps linearly onto ordinal FOIS across the full clinical spectrum, with a monotonic gradient from FDI-C 36.5 (FOIS 1, nothing by mouth) to 87.2 (FOIS 7, unrestricted oral diet). In that sense, FDI-C behaves as a continuous ruler of functional swallowing capacity, with near-perfect linearity in the proportional odds model (RCS p = 0.986).

Second, FDI-C achieves discriminative performance equivalent to clinician-rated severity for all outcomes (all p≥0.148), and correlates with it more strongly (ρ=−0.829) than the ordinal FEES severity score does (ρ=0.611). In this regard, a simple arithmetic mean approximates the holistic assessment that experienced clinicians perform intuitively.

Third, the clinical decision model demonstrates that the expert’s FOIS recommendation can be substantially reconstructed from three bedside-available variables. The full model including FDI-C, SPI and diagnosis achieves AUC 0.930 for FOIS≤3, with each predictor contributing incrementally (Table 5) and indicating excellent discrimination. Crucially, the direction of the diagnosis effect validates the clinical reasoning: at equivalent swallowing severity, stroke patients (acute dysphagia, compromised immune response, recovery expected) receive more restrictive diets than neurodegenerative patients (chronic adaptation, compensatory strategies established). This can be regarded as rational clinical practice formalized in a statistical model. That the diagnosis effect was most pronounced in the worst FDI-C quartile and vanished in the best quartile further supports this interpretation: when swallowing is severely impaired, clinical context drives the diet decision; when swallowing is intact, diagnosis is irrelevant for the degree of oral intake.

The implication is that FDI-C, together with functional status and diagnostic context, can approximate the FOIS level that an experienced clinician would recommend. This has the potential to standardize dysphagia severity assessment across settings, pending external validation.

### The mortality transition zone

The sigmoidal FDI-C–mortality relationship described above is more clinically informative than a linear gradient: If confirmed, the safe plateau would suggest that mild dysphagia does not contribute meaningfully to mortality risk. The apparent transition zone may identify a range where increasing swallowing impairment begins to matter: aspiration risk accumulates, nutritional status deteriorates, and clinical interventions become necessary. The apparent saturation below FDI-C ∼50 may reflect an intervention ceiling: patients with severe dysphagia are placed on maximal precautions (nil per os, tube feeding), so that further deterioration in swallowing physiology no longer translates into additional mortality.

With only 65 deaths and wide confidence intervals, particularly in the tails, this pattern should be considered hypothesis-generating. If confirmed in larger cohorts with sufficient events to test whether the threshold shifts by diagnosis or acuity, the transition zone could represent a suitable target range for clinical intervention and a natural enrichment criterion for therapeutic trials.

### FDI-C as a dual-use instrument

FDI-C has properties that position it for two complementary roles.

#### Clinical decision support

The linear FDI-C-FOIS mapping and the clinical decision model (AUC 0.930 for FOIS≤3) suggest that FDI-C could standardize how FEES findings are translated into diet recommendations. Unlike existing ordinal scores, which require subjective severity grading, FDI-C is computed directly from PAS and Yale ratings, i.e., the same standardized scales that FEES examiners already document. By making the link between swallowing physiology and diet recommendation explicit and reproducible, FDI-C may reduce inter-examiner variability in FEES interpretation - a known problem across centers [10]. Pending prospective validation, a conversion chart between FDI-C ranges and recommended FOIS levels (or ranges), stratified by stroke/non-stroke cohorts could serve as a practical decision aid. Also, the FDI would facilitate tracking the recovery trajectories of patients across centers (acute care - rehabilitation facilities - outpatient clinics) in the same way.

#### Research endpoint

FDI-C produces 387 unique values with confirmed linearity for the primary outcome (HAP: RCS p=0.60). By the asymptotic relative efficiency framework [27], categorizing a continuous variable into 5 ordinal levels requires approximately 10-15% more subjects. When the superior discrimination of FDI-C (which will further reduce unexplained variance in trial data) is additionally considered, the combined effect translates to approximately 15-22% fewer subjects required to detect a given effect size. FEDSS has been used as the primary endpoint in at least two published RCTs [37,38]; FDI could serve the same role with greater statistical efficiency.

The equal weighting of FDI-S and FDI-E deserves comment. Regression-derived optimal weights for FDI-S ranged from 0.51 (mortality) to 0.77 (FOIS), reflecting outcome-specific differences in the relative importance of safety versus efficiency. However, the AUC differences between optimally weighted and equally weighted composites were negligible for the distally measured outcomes and no DeLong comparison reached significance (all p > 0.5). Bootstrap 95% confidence intervals for the AUC-maximizing weight included 0.50 for three of four outcomes. The exception was FOIS, where the aspiration-dominated PAS component naturally dominates; however, this is the outcome most affected by the circularity limitation noted above and therefore least suitable for driving weight optimization. Equal weighting thus represents a principled, outcome-agnostic default that avoids overfitting (Online Resource 3).

That a single score can serve both roles simultaneously - guiding clinical decisions at the bedside while providing a continuous, bias-resilient outcome measure for research - is unusual in dysphagia assessment. Existing scores tend to be optimized for one purpose or the other. FDI-C bridges this gap by being simple enough for clinical use and precise enough for parametric analysis. It is also fully compatible with the integrated FEES report proposed by Dziewas and colleagues [4], which structures the clinical workflow from salient findings through severity grading and phenotyping to individualized treatment recommendations. FDI-C could serve as the central quantitative severity metric within this framework (Step 3), replacing the ordinal FEES dysphagia score with a continuous, bias-resilient alternative while preserving the framework’s clinical logic.

### Limitations

First, this is a single-center study. While the temporal replication across two cohorts separated by a multi-year gap provides stronger evidence than a single-cohort analysis, both cohorts share the same protocol, clinical team, and patient population. The FDI-C-FOIS mapping in particular may reflect institutional practice patterns. External validation in neurological and non-neurological cohorts is a logical and necessary next step; collaborations with external centers are ongoing.

Cross-center applicability also depends on consistency classification systems. The International Dysphagia Diet Standardisation Initiative (IDDSI) framework is increasingly adopted internationally [39]. FDI computation is agnostic to the specific consistency taxonomy (it averages across whatever consistencies are tested) but the specific FDI-C-FOIS mapping may vary with institutional consistency protocols and IDDSI levels used.

Second, the Cohort 2 IPW estimates for the thickened liquid branch were unstable (positivity violation at 35% testing rate, weights up to 39.3). However, the sensitivity analysis confirmed that the Naive-IPW Delta remained <1%, and Cohort 1’s well-behaved IPW (range 0.20-5.45) provides independent confirmation.

Third, the FOIS outcome is subject to some circularity: the FEES examiner is typically the same clinician who recommends FOIS. This circularity is inherent to the clinical workflow: FDI-C captures the swallowing information that informs the FOIS decision, so high correspondence is expected by design. Still, the distally measured outcomes (HAP, mortality) provide independent validation unaffected by this circularity.

Fourth, we operationalized our primary respiratory outcome as hospital-acquired pneumonia (HAP) using ICD-10 J69 coding during the inpatient stay. This is a pragmatic but imperfect proxy: J69 captures aspiration pneumonitis, bacterial pneumonia of presumed aspiration origin, and chemical pneumonitis, and retrospective coding cannot distinguish these entities; coding accuracy is also known to be variable across institutions. We therefore report HAP rather than "aspiration pneumonia" to avoid overstating the etiological attribution.

Fifth, FDI inherits PAS and Yale psychometric limitations. However, by averaging across consistencies, FDI mathematically reduces measurement error (Spearman-Brown effect), yielding expected composite reliability exceeding DIGEST-FEES from k≥2 onward.

Sixth, the mortality transition zone is based on just 65 events and therefore should be considered hypothesis-generating only. Confirmation in larger cohorts, ideally with sufficient events to test whether the threshold shifts by neurological diagnosis, is needed.

Seventh, we present no responsiveness-to-change data yet. The continuous nature of FDI-C suggests sensitivity to longitudinal change, but this requires demonstration.

Eighth, FDI operates on the clinician-documented PAS per consistency, typically recorded as the worst PAS across three to four swallow trials of that consistency [40]. By the same order-statistics logic that motivates the between-consistency averaging in FDI, this within-consistency maximum convention inflates per-consistency scores relative to a trial-level mean. Because the convention is uniform across all PAS-based FEES scores, it does not differentially disadvantage FDI; however, a trial-level FDI variant averaging all documented swallow trials rather than consistency summaries would be even more bias-resilient and is a logical extension pending routine trial-level documentation.

Ninth, although the FDI framework is conceptually agnostic to the imaging modality and PAS and Yale scores are equally applicable to videofluoroscopy (VFS), we developed and validated FDI exclusively in FEES cohorts. VFS differs in consistency protocols, trial structure, and rating conditions; whether the FDI framework, particularly the FDI-C-FOIS mapping and the sigmoidal FDI-C-mortality relationship, generalizes to VFS datasets remains to be investigated.

### Conclusions

The FEES Dysphagia Index, combining swallowing safety (PAS) and efficiency (Yale residue) via simple averaging, is a bias-resilient, bedside-calculable continuous score that captures the swallowing physiology component of expert clinical judgment. Derived in 1,257 patients and temporally replicated in 1,686, it produces 387 unique values with confirmed interval-scale properties and maps linearly onto clinician-assigned oral intake levels. Combined with functional status and diagnostic context, FDI-C reconstructs the expert’s diet recommendation with high accuracy and reliably predicts HAP with AUC 0.81. The sigmoidal relationship with mortality suggests a clinically meaningful transition zone that may inform enrichment strategies for therapeutic trials. These properties position FDI-C as both a clinical decision support tool and a continuous endpoint for dysphagia research, pending external validation.

## Data Availability

The dataset is not publicly available due to patient privacy regulations. Anonymized summary data and analysis code are available from the corresponding author upon reasonable request.

## Acknowledgements

The authors thank the FEES team at the Department of Neurology, RWTH Aachen University Hospital, for their commitment to standardized documentation of swallowing assessments.

## Statements and Declarations

### Ethics approval and consent to participate

This study was approved by the ethics committee of RWTH Aachen University (EK 088/17) and was performed in accordance with the ethical standards laid down in the 1964 Declaration of Helsinki and its later amendments. Individual informed consent was waived by the ethics committee due to the retrospective design and the exclusive use of routinely collected, pseudonymized clinical data.

### Competing interests

The authors declare that they have no competing interests.

### Funding

No external funding was received for this study.

### Authors’ contributions

CJW: Conceptualization, methodology, data curation, formal analysis, writing - original draft. ESG: Conceptualization, writing - review & editing. BM: Data curation, investigation, writing - review & editing. TM: Data curation, investigation, writing - review & editing. JP: Data curation, writing - review & editing. JBS: Supervision, writing - review & editing. BSW: Methodology, investigation, writing - review & editing. All authors read and approved the final manuscript.

## ONLINE RESOURCES

### Online Resource 1 — IPW diagnostics

Branch-specific propensity model details for both cohorts.

#### Cohort 1

Three branches (puree -> thin liquid, thin liquid -> solid, thin liquid -> thickened liquid). Baseline covariates at all branches: age, sex, HFRS, SPI, diagnosis. PAS scores from preceding consistencies included at all branches.

| Branch | n | Propensity<br>range | Weight<br>range | Max weight | Weights ><br>10 |
| --- | --- | --- | --- | --- | --- |
| Puree -><br>Thin liquid | 1,145 | 0.72-0.99 | 0.20-2.83 | 2.83 | 0 |
| Thin liquid<br>-> Solid | 1,062 | 0.42-0.96 | 0.37-5.45 | 5.45 | 0 |
| Thin liquid<br>-><br>Thickened<br>liquid | 1,062 | 0.18-0.78 | 0.43-4.12 | 4.12 | 0 |

Stabilized weight summary: mean 1.00 (SD 0.87), median 0.61, range 0.20-5.45.

#### Cohort 2

Three branches (puree -> liquid, liquid -> solid, liquid -> thickened liquid). Baseline covariates: age, sex, HFRS, SPI, diagnosis. PAS from preceding consistencies at all branches. Yale residue scores included only where likelihood ratio (LR) tests confirmed predictive relevance.

| Branch | n | Yale<br>include<br>d? | LR test<br>p | Propen<br>sity<br>range | Weight<br>range | Max<br>weight | Weights<br>> 10 |
| --- | --- | --- | --- | --- | --- | --- | --- |
| Puree<br>-><br>Liquid | 1,539 | No | 0.231 | 0.68-0.99 | 0.22-3.10 | 3.10 | 0 |
| Liquid<br>-> Solid | 1,480 | Yes | < 0.001 | 0.40-0.97 | 0.35-5.48 | 5.48 | 0 |
| Liquid<br>-><br>Thicken<br>ed<br>liquid | 1,480 | No (a<br>priori) | - | 0.08-0.72 | 0.47-39.3 | 39.3 | 175 |

The thickened liquid branch showed a positivity violation at a 35% testing rate, resulting in unstable weights. Propensity scores were truncated at the 1st and 99th percentiles.

### Online Resource 2 — Prolonged LOS results (Cohort 2)

Prolonged LOS defined as > median (16 days), prevalence 49.6%.

| Score | Prolonged LOS AUC (95% CI) |
| --- | --- |
| FDI-C | 0.614 (0.586-0.641) |
| FDI-S | 0.603 (0.575-0.631) |
| FDI-E | 0.596 (0.568-0.624) |
| Worst PAS | 0.559 (0.531-0.587) |
| Ordinal FEES severity score | 0.582 (0.556-0.607) |
| Clinician-rated severity | 0.629 (0.604-0.655) |
| FDI-C + SPI (clinical model) | 0.701 (0.673-0.728) |

DeLong comparisons: FDI-C vs Worst PAS: delta-AUC = +0.055, p = 0.006. FDI-C vs Clinician-rated severity: delta-AUC = -0.016, p = 0.419. RCS non-linearity p = 0.006.

### Online Resource 3 — FDI-S/FDI-E weighting sensitivity analysis

Sensitivity analysis examining whether alternative weightings of FDI-S (Safety) and FDI-E (Efficiency) improve the predictive performance of FDI-C over the equal-weight specification (FDI-C = 0.5 x FDI-S + 0.5 x FDI-E). Analyses performed in Cohort 2 (N = 1,557 with both FDI-S and FDI-E available).

#### Panel A: Regression-derived optimal weights

For each outcome, a logistic regression model was fitted with standardized FDI-S and FDI-E as separate predictors. The regression-derived weight for FDI-S was computed as w_S = |beta_S| / (|beta_S| + |beta_E|).

| Outcome | beta_S<br>(std) | beta_E<br>(std) | Ratio | w_S (regr) | w_E (regr) |
| --- | --- | --- | --- | --- | --- |
| HAP (J69) | -0.499 | -0.268 | 1.86 | 0.651 | 0.349 |
| Mortality | -0.338 | -0.328 | 1.03 | 0.508 | 0.492 |
| Prolonged<br>LOS | -0.291 | -0.214 | 1.36 | 0.577 | 0.423 |
| FOIS <= 3 | -1.987 | -0.597 | 3.33 | 0.769 | 0.231 |
| **Mean** |  |  |  | **0.626** | **0.374** |

Ratio = $|\text{beta}_S| / |\text{beta}_E|$ .

All beta coefficients from logistic regression with standardized predictors (both p < 0.05 for all outcomes).

#### Panel B: AUC across weight grid

FDI-C(w) = w x FDI-S + (1 - w) x FDI-E, with w from 0.00 to 1.00 (step 0.10).

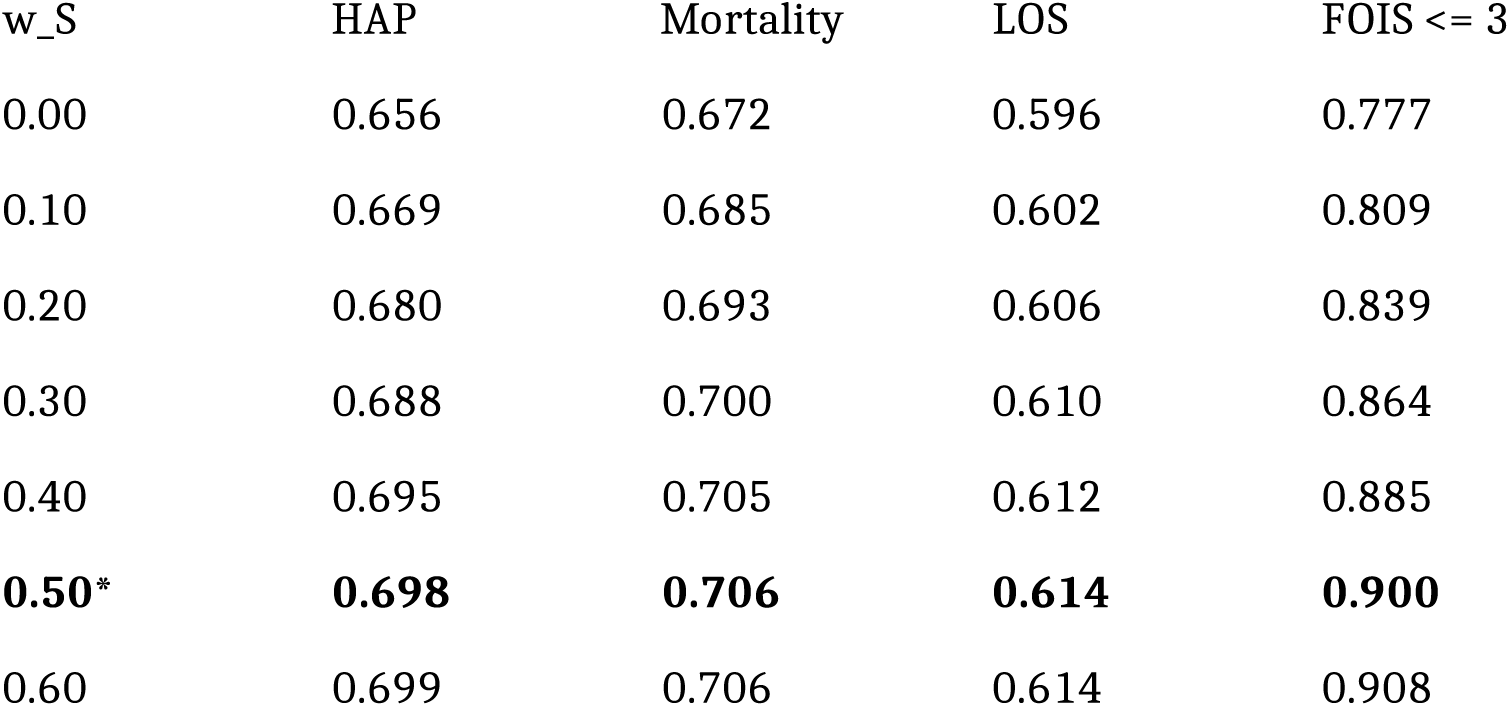
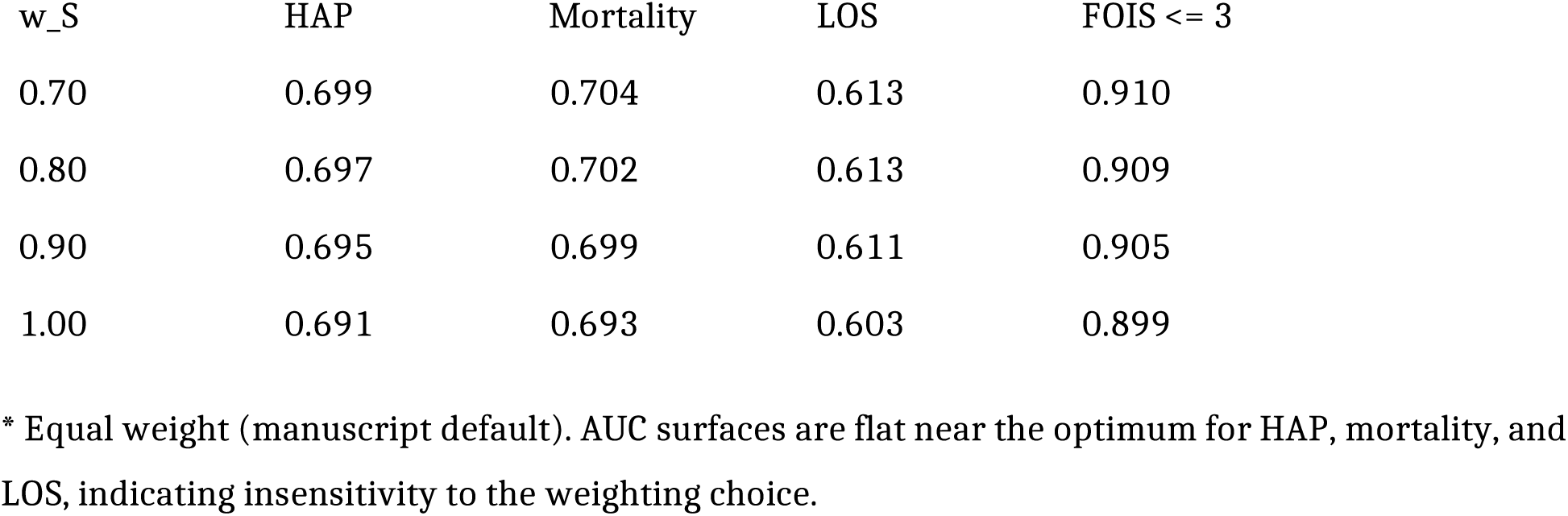

| w_S | HAP | Mortality | LOS | FOIS <= 3 |
| --- | --- | --- | --- | --- |
| 0.00 | 0.656 | 0.672 | 0.596 | 0.777 |
| 0.10 | 0.669 | 0.685 | 0.602 | 0.809 |
| 0.20 | 0.680 | 0.693 | 0.606 | 0.839 |
| 0.30 | 0.688 | 0.700 | 0.610 | 0.864 |
| 0.40 | 0.695 | 0.705 | 0.612 | 0.885 |
| **0.50\*** | **0.698** | **0.706** | **0.614** | **0.900** |
| 0.60 | 0.699 | 0.706 | 0.614 | 0.908 |
| 0.70 | 0.699 | 0.704 | 0.613 | 0.910 |
| 0.80 | 0.697 | 0.702 | 0.613 | 0.909 |
| 0.90 | 0.695 | 0.699 | 0.611 | 0.905 |
| 1.00 | 0.691 | 0.693 | 0.603 | 0.899 |

\* Equal weight (manuscript default). AUC surfaces are flat near the optimum for HAP, mortality, and LOS, indicating insensitivity to the weighting choice.

#### Panel C: AUC-maximizing weights, DeLong comparisons, and bootstrap CIs

| Outcome | Best w_S | AUC(best<br>) | AUC(0.50<br>) | delta-AUC | DeLong p | Bootstra<br>p 95% CI |
| --- | --- | --- | --- | --- | --- | --- |
| HAP<br>(J69) | 0.65 | 0.699 | 0.698 | +0.001 | 0.797 | [0.40,<br>1.00] |
| Mortality | 0.50 | 0.706 | 0.706 | 0.000 | 0.507 | [0.15,<br>0.95] |
| Prolonge<br>d LOS | 0.55 | 0.614 | 0.614 | +0.000 | 0.955 | [0.25,<br>0.95] |
| FOIS <= 3 | 0.75 | 0.910 | 0.900 | +0.010 | 0.007 | [0.65,<br>0.80] |

DeLong tests compare AUC-maximizing weighted FDI-C (from Panel B grid search) against equal-weighted FDI-C. Bootstrap 95% CIs based on 1,000 resamples. The bootstrap CI includes 0.50 for three of four outcomes. The DeLong test for FOIS is significant (p = 0.007), but FOIS is the outcome most affected by the circularity limitation (see main text). FDI-S and FDI-E correlation: Pearson r = 0.538.

#### Interpretation

Regression-derived optimal weights are outcome-specific (w_S range: 0.51-0.77), confirming that no single alternative weighting uniformly outperforms equal weighting. For the distally measured, clinically meaningful outcomes (aspiration pneumonia, mortality), AUC differences between optimal and equal weighting are negligible (delta-AUC <= 0.001) and statistically non-significant. The equal-weight specification is justified as an outcome-agnostic, overfitting-resistant default that maintains clinical interpretability.

### Online Resource 4 — Derivation/Validation split AUCs (Cohort 2)

Stratified 60/40 random split (derivation n = 1,011, validation n = 675).

| Score | Outcome | Derivation<br>AUC | Validation<br>AUC | delta-AUC | Stability |
| --- | --- | --- | --- | --- | --- |
| FDI-C | Aspiration pneumonia | 0.709 | 0.687 | -0.022 | Moderate |
| FDI-C | Mortality | 0.703 | 0.709 | +0.006 | Stable |
| FDI-C | FOIS <= 3 | 0.902 | 0.896 | -0.006 | Stable |
| FDI-S | Aspiration pneumonia | 0.700 | 0.675 | -0.025 | Moderate |
| FDI-S | Mortality | 0.696 | 0.693 | -0.003 | Stable |
| FDI-S | FOIS <= 3 | 0.901 | 0.890 | -0.011 | Stable |
| FDI-E | Aspiration pneumonia | 0.660 | 0.653 | -0.007 | Stable |
| FDI-E | Mortality | 0.654 | 0.697 | +0.043 | Moderate |
| FDI-E | FOIS <= 3 | 0.790 | 0.757 | -0.033 | Moderate |

Stability classification: |delta-AUC| < 0.02 = stable, 0.02-0.05 = moderate, > 0.05 = unstable.

FDI-C remained significantly superior to Worst PAS in the validation set for aspiration pneumonia (p = 0.007) and FOIS <= 3 (p < 0.001), and equivalent to clinician-rated severity (all p >= 0.243).

### Online Resource 5 — Inter-consistency correlations

#### Cohort 1 (PAS, Spearman)

| Pair | rho | p | n |
| --- | --- | --- | --- |
| Puree vs Thin liquid | 0.329 | < 0.001 | 1,062 |
| Puree vs Thickened liquid | 0.316 | < 0.001 | 521 |
| Puree vs Solid | 0.209 | < 0.001 | 741 |
| Thin liquid vs Thickened liquid | 0.320 | < 0.001 | 532 |
| Thin liquid vs Solid | 0.135 | < 0.001 | 763 |
| Thickened liquid vs Solid | 0.183 | 0.003 | 350 |

#### Cohort 2 (PAS, Spearman)

| Pair | rho | p | n |
| --- | --- | --- | --- |
| Puree vs Liquid | 0.325 | < 0.001 | 1,480 |
| Puree vs Thickened liquid | 0.243 | < 0.001 | 592 |
| Puree vs Solid | 0.141 | < 0.001 | 1,136 |
| Liquid vs Thickened liquid | 0.150 | < 0.001 | 589 |
| Liquid vs Solid | 0.130 | < 0.001 | 1,142 |
| Thickened liquid vs Solid | 0.151 | < 0.001 | 417 |

#### Cohort 2 (Yale combined, Spearman)

| Pair | rho | p | n |
| --- | --- | --- | --- |
| Puree vs Liquid | 0.527 | < 0.001 | 1,472 |
| Puree vs Thickened liquid | 0.472 | < 0.001 | 589 |
| Puree vs Solid | 0.539 | < 0.001 | 1,145 |
| Liquid vs<br>Thickened liquid | 0.599 | < 0.001 | 583 |
| Liquid vs Solid | 0.377 | < 0.001 | 1,141 |
| Thickened liquid<br>vs Solid | 0.446 | < 0.001 | 418 |

PAS correlations are moderate (rho = 0.13-0.33), confirming that swallowing safety with one consistency does not reliably predict safety with others. Yale correlations are higher (rho = 0.38-0.60), consistent with pharyngeal residue being a more stable trait.

### Online Resource 6 — Sensitivity analysis: IPW with vs without thickened liquid branch (Cohort 2)

#### Three-cohort bias comparison under both IPW specifications

| Score | Subcohort | IPW full model | IPW no-thick model |
| --- | --- | --- | --- |
| FDI-S | CC mean (n = 412) | 66.97 | 66.97 |
|  | SCD mean (n = 1,480) | 67.24 | 67.24 |
|  | IPW reference | 65.70 | 74.13 |
|  | Naive-IPW Delta | 0.5% | 0.4% |
| FDI-E | CC mean (n = 412) | 61.45 | 61.45 |
|  | SCD mean (n = 1,480) | 65.04 | 65.04 |
|  | IPW reference | 61.90 | 65.58 |
|  | Naive-IPW Delta | 0.6% | 0.3% |
| FDI-C | CC mean (n = 412) | 64.21 | 64.21 |
|  | SCD mean (n = 1,480) | 66.14 | 66.14 |
|  | IPW reference | 63.81 | 69.73 |
|  | Naive-IPW Delta | 0.6% | 0.4% |
| Worst PAS | CC mean (n = 412) | 7.02 | 7.02 |
|  | SCD mean (n = 1,480) | 4.84 | 4.84 |
|  | IPW reference | 6.39 | 4.71 |

The IPW reference values diverge substantially between the two specifications (e.g., FDI-S: 65.70 vs 74.13, delta = 8.43 points), confirming that the exact IPW-corrected population mean is sensitive to modeling choices in the presence of positivity violations. However, the Naive-IPW Delta remains below 1% under both specifications for all FDI variants, demonstrating that FDI’s bias resilience is not dependent on any particular IPW model.

### Online Resource 7 — Yale weighting sensitivity analysis (Cohort 2)

#### Panel A: Pooled Yale-FOIS regression (cluster-robust, N = 4,755 observations from 1,555 patients)

| Predictor | Beta | p | Relative weight |
| --- | --- | --- | --- |
| Yale Valleculae | -0.211 | < 0.001 | 0.29 |
| Yale Sinus Piriformis | -0.527 | < 0.001 | 0.71 |

R-squared = 0.184. Cluster-robust standard errors (patient-level clustering).

#### Panel B: Per-consistency Yale-FOIS regressions

R-squared = 0.184. Cluster-robust standard errors (patient-level clustering). Panel B: Per-consistency Yale-FOIS regressions
| Consistency | n | R-squared | Beta Vall (p) | Beta SP (p) | Weight Vall : SP |
| --- | --- | --- | --- | --- | --- |
| Puree | 1,467 | 0.179 | -0.139 (< 0.001) | -0.579 (< 0.001) | 0.19 : 0.81 |
| Liquid | 1,445 | 0.082 | -0.254 (0.002) | -0.435 (< 0.001) | 0.37 : 0.63 |
| Thickened liquid | 591 | 0.056 | -0.246 (0.002) | -0.101 (0.136) | 0.71 : 0.29 |
| Solid | 1,143 | 0.125 | -0.251 (< 0.001) | -0.198 (< 0.001) | 0.56 : 0.44 |

Relative weights vary across consistencies and reverse between thickened liquid (valleculae-dominant) and puree (pyriform sinus-dominant).

#### Panel C: FOIS correlation comparison (simple mean vs empirically weighted Yale)

| Consistency | rho (simple mean, FOIS) | rho (weighted, FOIS) | delta-rho |
| --- | --- | --- | --- |
| Puree | -0.495 | -0.478 | +0.017 |
| Liquid | -0.358 | -0.356 | +0.002 |
| Thickened liquid | -0.208 | -0.205 | +0.004 |
| Solid | -0.387 | -0.377 | +0.011 |

The simple mean achieves equal or marginally higher FOIS correlations across all consistencies.

#### Panel D: FDI-E variant comparison

| Variant | Mean (SD) | Pearson r |
| --- | --- | --- |
| FDI-E (simple mean) | 64.5 (21.3) | - |
| FDI-E (weighted, 0.29 x<br>Vall + 0.71 x SP) | 67.8 (21.6) | 0.9845 |

The two FDI-E variants are near-identical (r = 0.98). Because the relative weight of valleculae vs pyriform sinuses reverses between consistencies (Panel B), the pooled empirical weights do not generalize across testing protocols. The simple mean was retained for parsimony and generalizability.

### Online Resource 8 — STROBE checklist for cohort studies

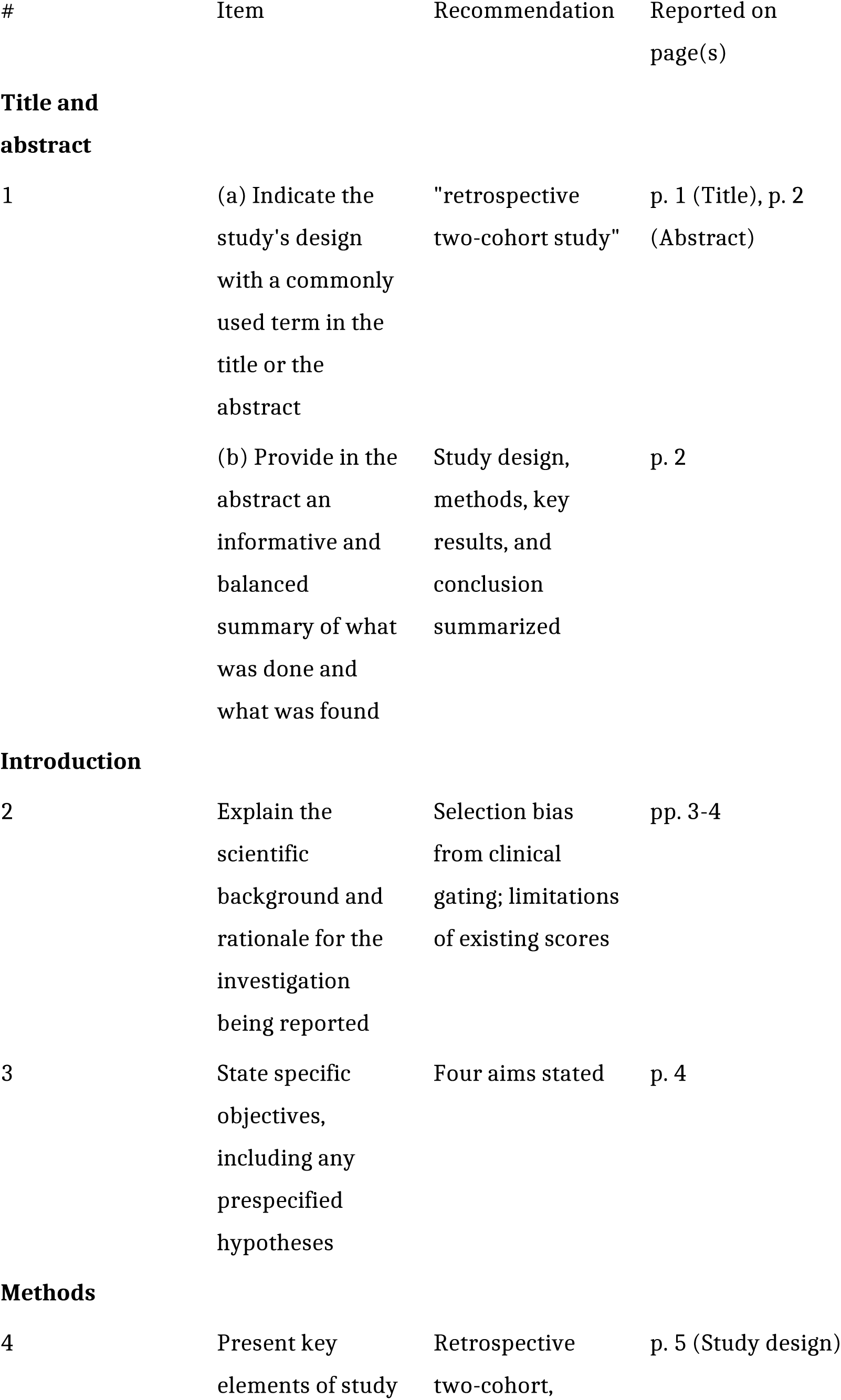
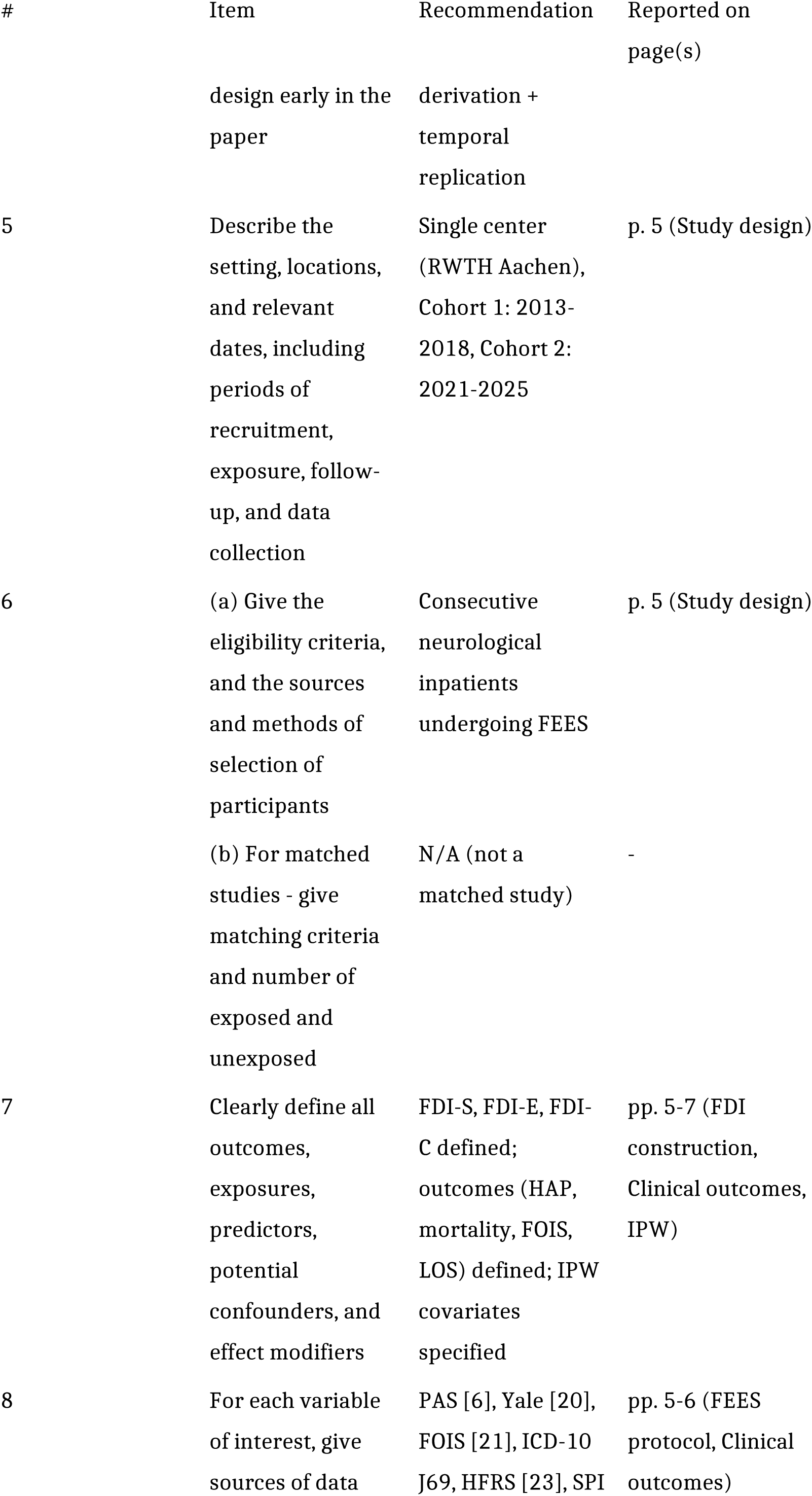
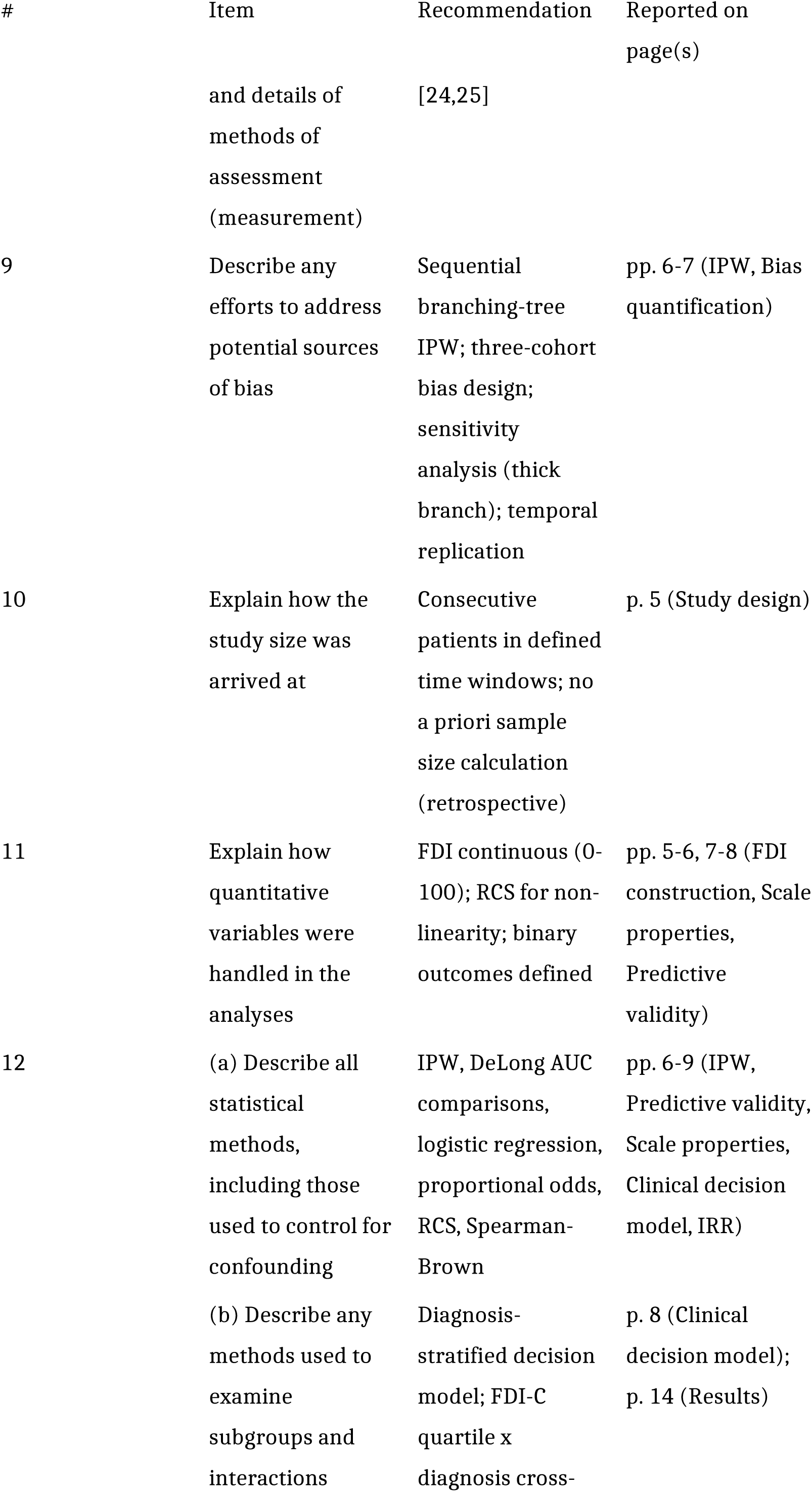
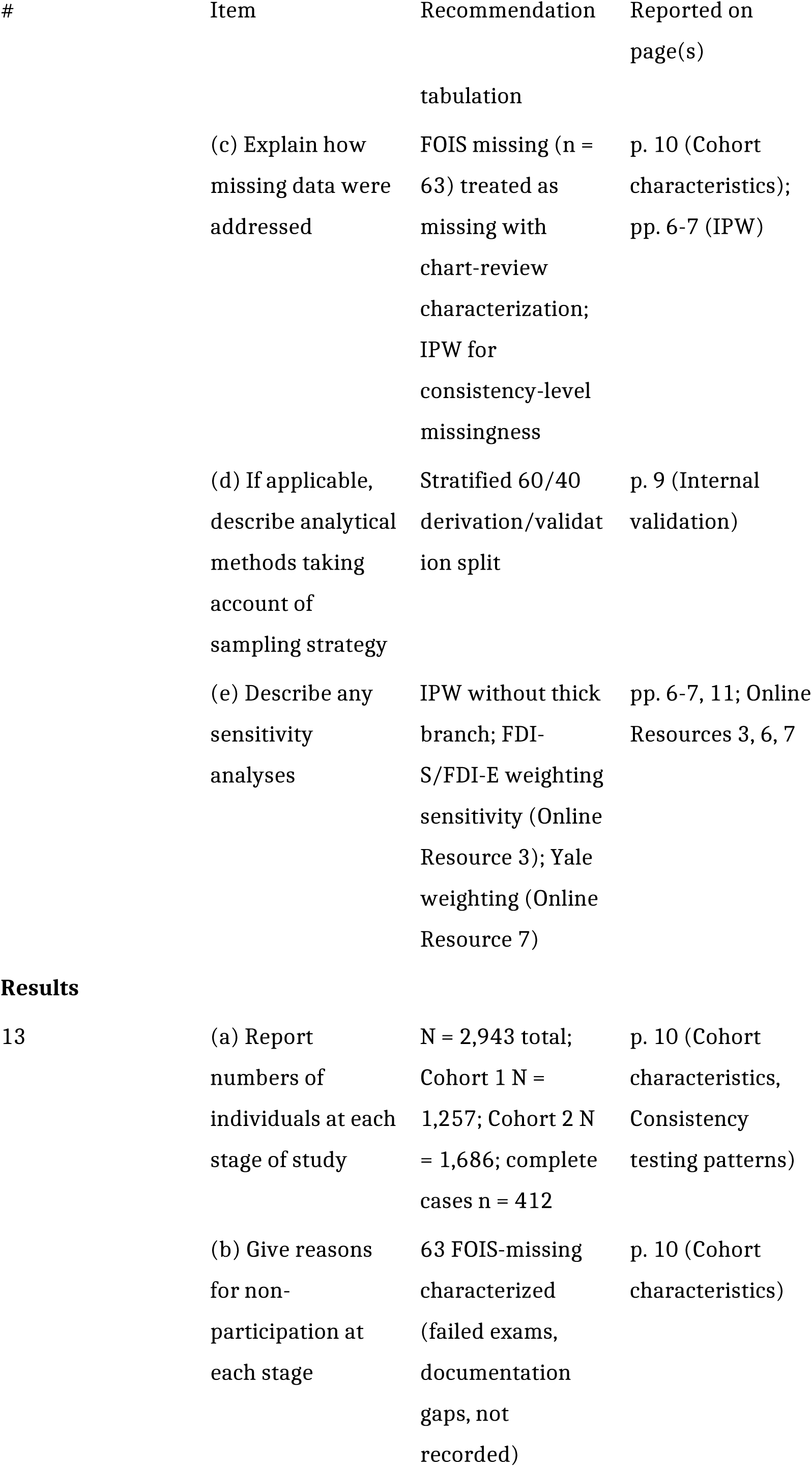
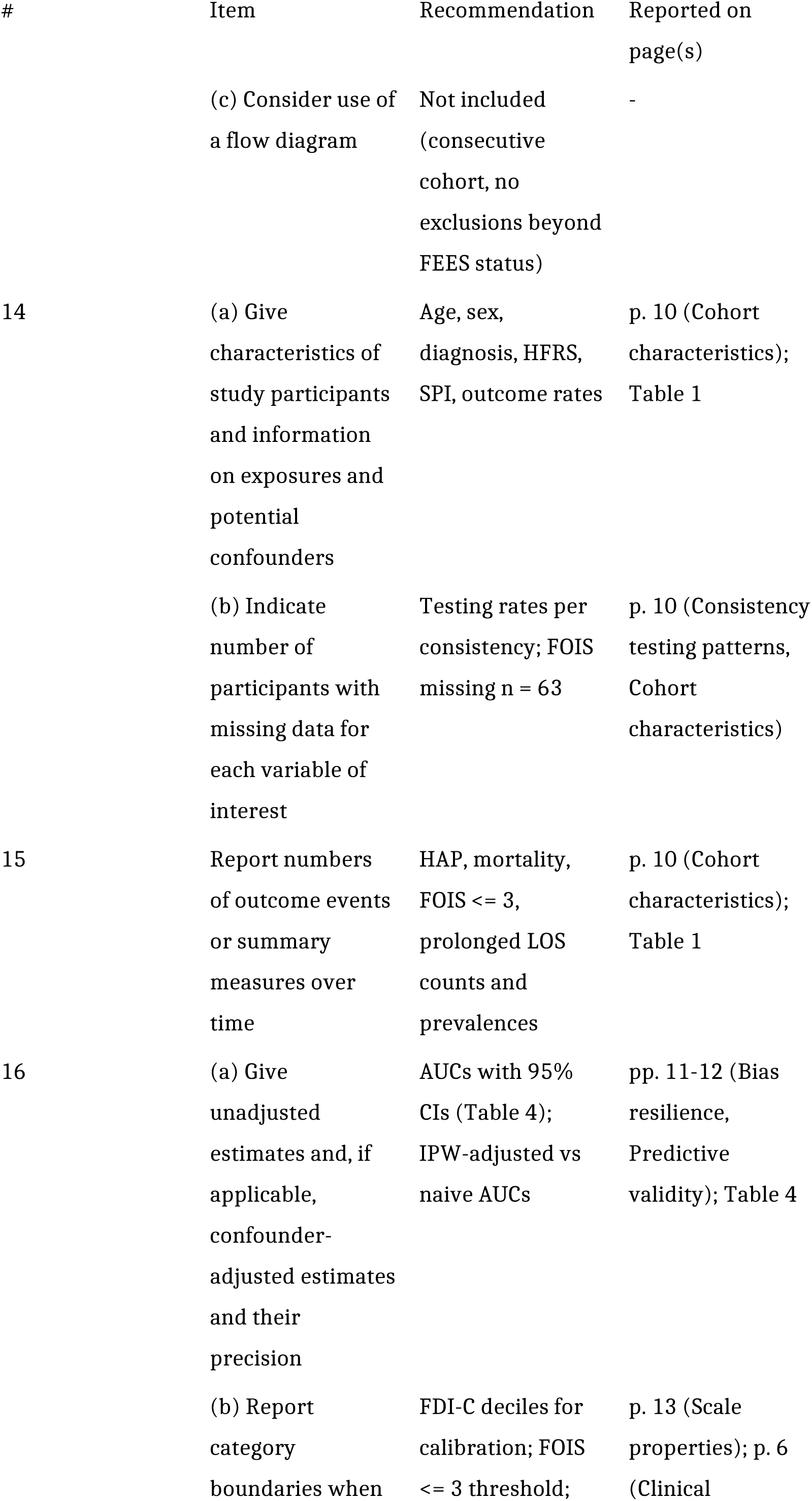
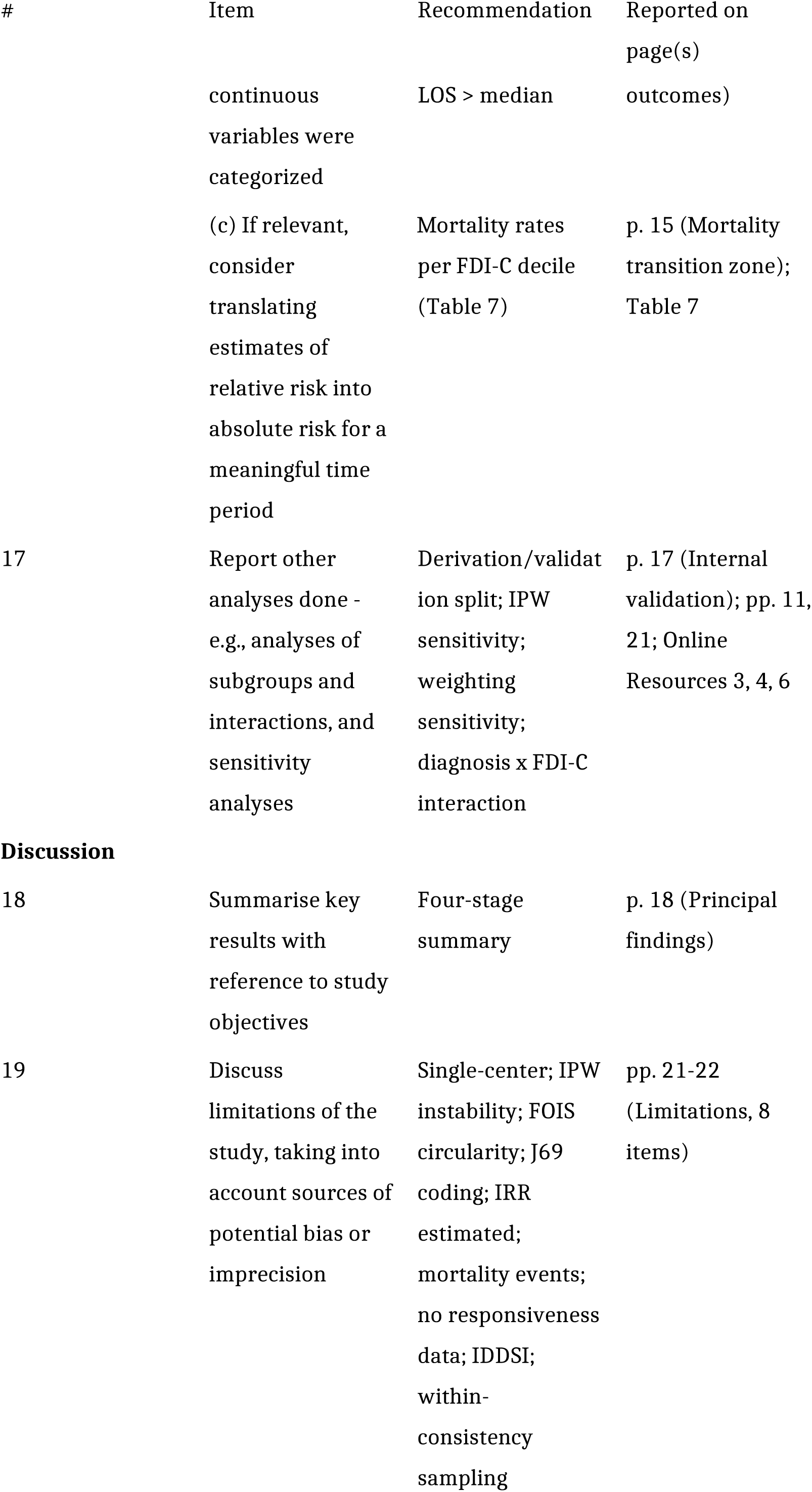
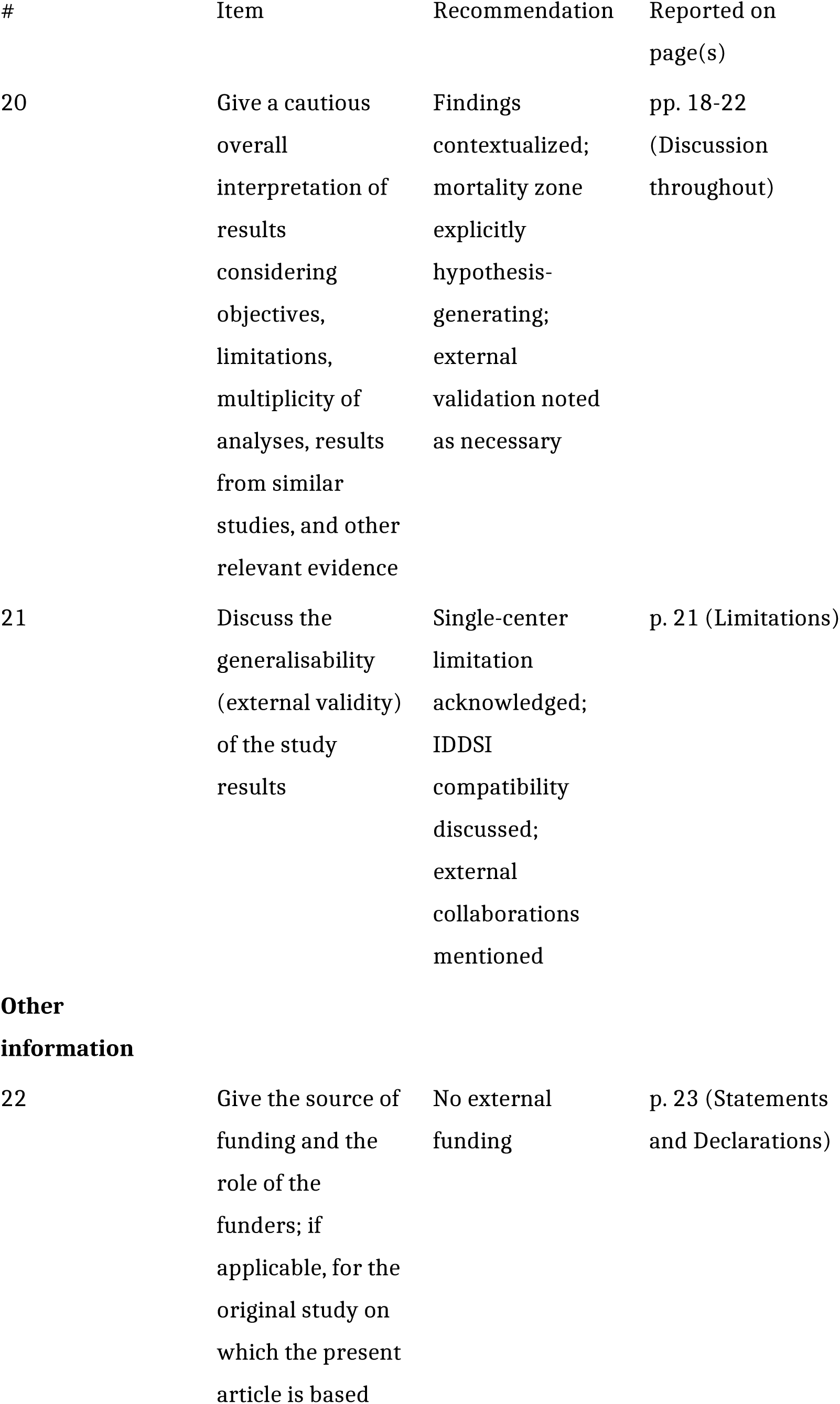

Reference: von Elm E, Altman DG, Egger M, Pocock SJ, Gotzsche PC, Vandenbroucke JP. The Strengthening the Reporting of Observational Studies in Epidemiology (STROBE) statement: guidelines for reporting observational studies. *Lancet* 2007;370:1453–1457.

## Notes

### Competing Interest Statement

The authors have declared no competing interest.

## References

[1] Langmore SE, Kenneth SMA, Olsen N. Fiberoptic endoscopic examination of swallowing safety: A new procedure. Dysphagia 1988;2:216–9. 10.1007/BF02414429.

[2] Langmore SE. Endoscopic evaluation of oral and pharyngeal phases of swallowing. GI Motility Online 2006. 10.1038/gimo28.

[3] Dziewas R, Glahn J, Helfer C, et al. Flexible endoscopic evaluation of swallowing (FEES) for neurogenic dysphagia: training curriculum of the German Society of Neurology and the German stroke society. BMC Med Educ 2016;16:70. 10.1186/s12909-016-0587-3.

[4] Dziewas R, Warnecke T, Labeit B, et al. Systematic approach to contextualize findings of flexible endoscopic evaluation of swallowing in neurogenic dysphagia– towards an integrated FEES report. Neurol Res Pract 2024;6:26. 10.1186/s42466-024-00321-8.

[5] Dziewas R, Michou E, Trapl-Grundschober M, et al. European Stroke Organisation and European Society for Swallowing Disorders guideline for the diagnosis and treatment of post-stroke dysphagia. Eur Stroke J 2021;6:LXXXIX–CXV. 10.1177/23969873211039721.

[6] Rosenbek JC, Robbins JA, Roecker EB, et al. A penetration-aspiration scale. Dysphagia 1996;11:93–8. 10.1007/BF00417897.

[7] Steele CM, Grace-Martin K. Reflections on Clinical and Statistical Use of the Penetration-Aspiration Scale. Dysphagia 2017;32:601–16. 10.1007/s00455-017-9809-z.

[8] Borders JC, Brates D. Use of the Penetration-Aspiration Scale in Dysphagia Research: A Systematic Review. Dysphagia 2020;35:583–97. 10.1007/s00455-019-10064-3.

[9] Borders JC, Steele CM. The effect of liquid consistency on penetration-aspiration: a Bayesian analysis of two large datasets. Front Rehabil Sci 2024;5. 10.3389/fresc.2024.1337971.

[10] Wehner A, Schumann-Werner B, Fimm B, et al. [Standards in the use of fiberoptic endoscopic evaluation of swallowing in Germany : A questionnaire survey]. Nervenarzt 2021. 10.1007/s00115-021-01127-8.

[11] Borders JC, Grande AA, Barbon CEA, et al. Effects of Statistical Practices for Longitudinal Group Comparison of the Penetration-Aspiration Scale on Power and Effect Size Estimation: A Monte Carlo Simulation Study. Dysphagia 2025;40:388–98. 10.1007/s00455-024-10738-7.

[12] Starmer HM, Arrese L, Langmore S, et al. Adaptation and Validation of the Dynamic Imaging Grade of Swallowing Toxicity for Flexible Endoscopic Evaluation of Swallowing: DIGEST-FEES. J Speech Lang Hear Res 2021;64:1802–10. 10.1044/2021_JSLHR-21-00014.

[13] Labeit B, Lapa S, Muhle P, et al. Validation of the DIGEST-FEES as a Global Outcome Measure for Pharyngeal Dysphagia in Parkinson’s Disease. Dysphagia 2023. 10.1007/s00455-023-10650-6.

[14] Warnecke T, Ritter MA, Kröger B, et al. Fiberoptic Endoscopic Dysphagia Severity Scale Predicts Outcome after Acute Stroke. CED 2009;28:283–9. 10.1159/000228711.

[15] Dziewas R, Warnecke T, Ölenberg S, et al. Towards a Basic Endoscopic Assessment of Swallowing in Acute Stroke – Development and Evaluation of a Simple Dysphagia Score. CED 2008;26:41–7. 10.1159/000135652.

[16] Warnecke T, Oelenberg S, Teismann I, et al. Endoscopic characteristics and levodopa responsiveness of swallowing function in progressive supranuclear palsy. Mov Disord 2010;25:1239–45. 10.1002/mds.23060.

[17] Schröder JB, Marian T, Muhle P, et al. Intubation, tracheostomy, and decannulation in patients with Guillain–Barré–syndrome—does dysphagia matter? Muscle and Nerve 2019;59:194–200. 10.1002/mus.26377.

[18] Warnecke T, Im S, Labeit B, et al. Detecting myasthenia gravis as a cause of unclear dysphagia with an endoscopic tensilon test. Ther Adv Neurol Disord 2021;14:17562864211035544. 10.1177/17562864211035544.

[19] Lapa S, Claus I, Reitz SC, et al. Effect of thalamic deep brain stimulation on swallowing in patients with essential tremor. Ann Clin Transl Neurol 2020;7:1174–80. 10.1002/acn3.51099.

[20] Neubauer PD, Rademaker AW, Leder SB. The Yale Pharyngeal Residue Severity Rating Scale: An Anatomically Defined and Image-Based Tool. Dysphagia 2015;30:521–8. 10.1007/s00455-015-9631-4.

[21] Crary MA, Mann GDC, Groher ME. Initial Psychometric Assessment of a Functional Oral Intake Scale for Dysphagia in Stroke Patients. Archives of Physical Medicine and Rehabilitation 2005;86:1516–20. 10.1016/j.apmr.2004.11.049.

[22] Austin PC, Stuart EA. Estimating the effect of treatment on binary outcomes using full matching on the propensity score. Stat Methods Med Res 2017;26:2505–25. 10.1177/0962280215601134.

[23] Gilbert T, Neuburger J, Kraindler J, et al. Development and validation of a Hospital Frailty Risk Score focusing on older people in acute care settings using electronic hospital records: an observational study. The Lancet 2018;391:1775–82. 10.1016/S0140-6736(18)30668-8.

[24] Koch D, Schuetz P, Haubitz S, et al. Improving the post-acute care discharge score (PACD) by adding patients’ self-care abilities: A prospective cohort study. PLoS One 2019;14:e0214194. 10.1371/journal.pone.0214194.

[25] Koch D, Kutz A, Haubitz S, et al. Association of functional status and hospital-acquired functional decline with 30-day outcomes in medical inpatients: A prospective cohort study. Appl Nurs Res 2020;54:151274. 10.1016/j.apnr.2020.151274.

[26] DeLong ER, DeLong DM, Clarke-Pearson DL. Comparing the areas under two or more correlated receiver operating characteristic curves: a nonparametric approach. Biometrics 1988;44:837–45.

[27] Royston P, Altman DG, Sauerbrei W. Dichotomizing continuous predictors in multiple regression: a bad idea. Stat Med 2006;25:127–41. 10.1002/sim.2331.

[28] Altman DG, Royston P. The cost of dichotomising continuous variables. BMJ 2006;332:1080.1. 10.1136/bmj.332.7549.1080.

[29] Harrell, FE. Regression Modeling Strategies: With Applications to Linear Models, Logistic and Ordinal Regression, and Survival Analysis. Cham: Springer International Publishing; 2015. 10.1007/978-3-319-19425-7.

[30] Peduzzi P, Concato J, Kemper E, et al. A simulation study of the number of events per variable in logistic regression analysis. Journal of Clinical Epidemiology 1996;49:1373–9. 10.1016/S0895-4356(96)00236-3.

[31] Nunnally JC, Bernstein IH. Psychometric theory. 3rd ed. New York: McGraw-Hill; 1994.

[32] Butler SG, Markley L, Sanders B, et al. Reliability of the penetration aspiration scale with flexible endoscopic evaluation of swallowing. Ann Otol Rhinol Laryngol 2015;124:480–3. 10.1177/0003489414566267.

[33] Pisegna JM, Borders JC, Kaneoka A, et al. Reliability of Untrained and Experienced Raters on FEES: Rating Overall Residue is a Simple Task. Dysphagia 2018;33:645–54. 10.1007/s00455-018-9883-x.

[34] R Core Team. R 2024.

[35] Robin X, Turck N, Hainard A, et al. pROC: an open-source package for R and S+ to analyze and compare ROC curves. BMC Bioinformatics 2011;12:77. 10.1186/1471-2105-12-77.

[36] David HA, Nagaraja HN. Order Statistics. 1st ed. Wiley; 2003. 10.1002/0471722162.

[37] Suntrup-Krueger S, Ringmaier C, Muhle P, et al. Randomized trial of transcranial direct current stimulation for poststroke dysphagia. Ann Neurol 2018;83:328–40. 10.1002/ana.25151.

[38] Suntrup-Krueger S, Labeit B, Von Itter J, et al. Treating postextubation dysphagia after stroke with pharyngeal electrical stimulation –insights from a randomized controlled pilot trial. Neurotherapeutics 2025;22:e00613. 10.1016/j.neurot.2025.e00613.

[39] Cichero JAY, Lam P, Steele CM, et al. Development of International Terminology and Definitions for Texture-Modified Foods and Thickened Fluids Used in Dysphagia Management: The IDDSI Framework. Dysphagia 2017;32:293–314. 10.1007/s00455-016-9758-y.

[40] Baijens LWJ, Speyer R, Pilz W, et al. FEES Protocol Derived Estimates of Sensitivity: Aspiration in Dysphagic Patients. Dysphagia 2014;29:583–90. 10.1007/s00455-014-9549-2.

